# Breast cancer: Emerging principles of metastasis, adjuvant and neoadjuvant treatment from cancer registry data

**DOI:** 10.1101/2020.12.01.20214551

**Authors:** Jutta Engel, Renate Eckel, Simone Schrodi, Kathrin Halfter, Gabriele Schubert-Fritschle, Dieter Hölzel

**Author notes:** Corresponding author: Prof. Dr. Dieter Hölzel, Munich Cancer Registry, D-81377 München, Germany.

## Abstract

**Background:** Growing primary breast cancers (PT) can initiate local (LR), regional (pLN), and distant metastases (MET). Characteristics of these progressions such as initiation, frequency, growth duration and treatment success describe principles of these processes. They are bottlenecks through which scientific and molecular biological concepts and hypotheses must fit.

**Methods:** Population-based data from the Munich Cancer Registry over 4 time periods since 1978 with the most important prognostic factors and an up to date follow-up are analyzed. With 66.818 patients, reliable data are obtained on initiation on METs, growth time und survival even in small subgroups. Together with results of clinical trials on prevention and adjuvant treatment (AT) principles for tumor growth, MET process and AT are derived.

**Results:** The median growth periods for PT/ MET/LR/pLN result in 12.5/8.8/5/3.5 years. Even if 30% of METs only appear after 10 years of MET-free time, a delayed initiation or cascade like initiation of METs, e.g. from pLNs cannot be derived from the data. That is an immediate MET initiation principle by PT. The growth rate of the PT can vary by a factor of 10 or more and can be transferred to the MET. Nevertheless, the relation of the growth times PT/MET results in a less varying value of 1.4. Principles of AT are the 50% eradication of 1st and 2ndPTs, the selective and partial eradication of bone and lung METs with successful ATs, which cannot be improved by extending the duration of ATs. These principles reveal, among other things, that there is no rationale for the accepted for long-term endocrine ATs, breast cancer risk by hormone replacement therapies, or cascading initiation of METs.

**Conclusion:** A paradigm with ten principles for the MET process and ATs can be derived from real world data and clinical trials. The principles show limits and opportunities for innovation also through alternative interpretations of well-known studies. The outlined MET process should be generalizable to all solid tumors.

## Introduction

Biomedical research regularly increases the complexity of cancer. More and more details are emerging on the association with risk factors, the steps involved in carcinogenesis, and processes such as clonal evolution that ultimately result in genetically heterogeneous PTs.^1-3^ Steps leading to metastasis or the MET-process (MET-P), are also becoming increasingly differentiated.^4,5^ By definition secondary metastases (sMET) are referred to as local recurrences (LRs), positive lymph nodes (pLNs), and distant METs. Primary and secondary BCs (1stPT/2ndPT) are included because these may also be prevented and have the same risk of initiating sMETs.

Even though the molecular processes involved in the disease course of PTs and sMETs are becoming increasingly complex, they can still be described with the known parameters and a few principles of METs and their treatment. The aim of this article is to elucidate initiation, growth, survival, and treatment effects of BC and sMET with real world data from the Munich Cancer Registry (MCR) which are basic conditions, bottlenecks through which molecular hypotheses and scientific terms for prognosis and prediction have to go.

## Methods

The Munich Cancer registry (MCR) has data starting from 1978. It has been population-based for the currently underlying 4.9 million population since 1998, and is included in Cancer Incidence of Five Continents. Reliable data on changing adjuvant treatments (AT) and on locoregional disease manifestations from pathology reports such as hormone receptor status, tumor diameter (TD), number of pLN, Ki67 - since 2010 - and contralateral PT are available. All death certificates from the region are included and provide an up-to-date complete follow-up.

In the case of cancer-related death, approximately 70% of METs were recorded. Four time intervals have been distinguished since 1978 and the time trends of the successful ATs and changing progressions are analyzed. Despite missing values, it is robust data with 66.818 patients for the analysis of even small subgroups.

Kaplan-Meier curves for the relative survival from diagnosis, for survival up to and after MET and with distributions of the MET-free survival time describe the relationships with prognostic and predictive factors. The relative survival is an estimate for tumor-specific survival and is calculated by dividing the overall survival after diagnosis by the survival observed in the general population with comparable age distribution.

METs account for the great majority of cancer-associated deaths, why this complex process needs to be better understood. With every millimeter of PT growth, further METs are initiated. Three growth phases are to be estimated, the time from occult MET growth to PT diagnosis, the MET-free time up to MET detection and survival afterwards. The distribution of the MET-free survival times excludes the growth time in the occult phase. However, MET growth can be estimated using twice the median MET free time because MET growth is not expected to change with PT diagnosis.

So far, modeling has seldom been used in medicine to elucidate relationships. Processes are modeled with the distribution functions for the incidence of PTs and METs, the growth times and eradication rates. The effect of HRT with growth acceleration of prevalent PTs and a risk of new PTs or the long-term endocrine AT with a preventive and adjuvant effect are examples. Statistical analyses were performed by using SAS V 9.4 and R V 3.1.3.

## Results

### Initiation of primary tumors and secondary foci

Growing and evolutionarily developing PTs may have already disseminated DNA (ctDNA) and circulating tumor cells (TC) to form local or distant small foci through alterations in the tumor microenvironment.^6,7^ Several cell types and signaling molecules are involved in promoting the epithelial-mesenchymal transition that allow TCs to disseminate. PTs can achieve MET-competence starting from about 1 mm TD^4^ and initially a first MET-competent TC appears among many disseminated TCs.^8^ They may remain local or may spread through lymphatic or hematogenous dissemination and initiate LRs, pLNs, and METs, which grow in parallel and are discovered at the earliest with the PT diagnosis. Initiation and growth of PT and these sMETs are shown in Fig.1. In Fig.2A, possible sources and pathways of the initiating TCs are outlined and some results for pT1c- and pT2-PTs are arranged in Fig.2B. Growing PTs are associated with worsening prognosis (Fig.2C). Until R0-resection, all TCs that can initiate sMETs are disseminated. Only dormancy could delay the onset. In the form of a liquid biopsy, circulating TCs and ctDNA can be detected when diagnosing a BC, although they are very rare and the sensitivity and specificity of the procedure is moderate.^9-11^

**Figure 1.**
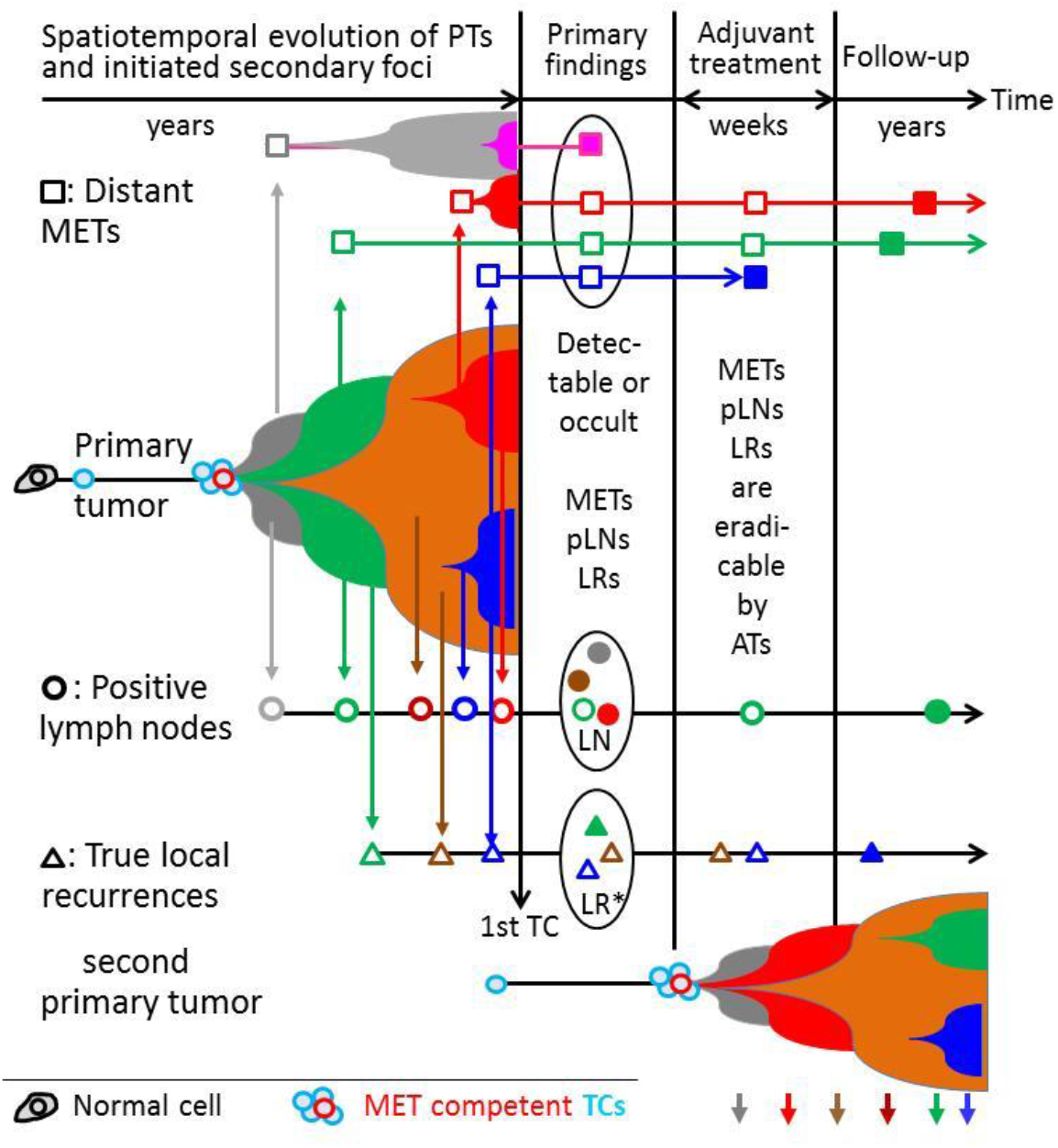
Initiation and growth of a PT and secondary foci. Growing PTs can initiate LRs, pLNs and METs with different gene signature. They can be diagnosed synchronously with PTs (filled symbols), remain occult, will be eradicated by ATs or occur in the course of disease. The article and this figure were inspired by LR Yates et al (2017).^3^

**Figure 2:**
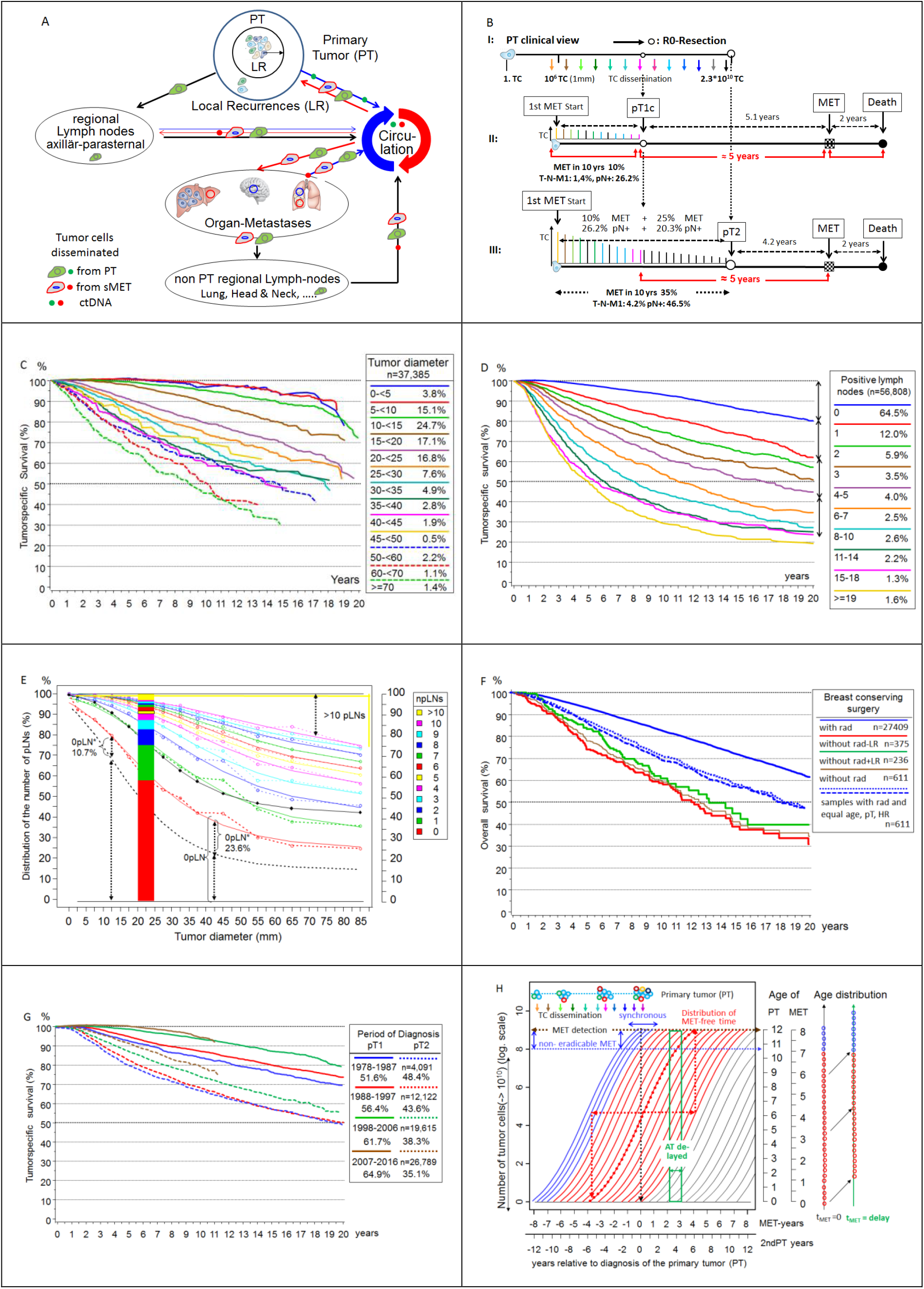
2A – 2H combined (A) Possible sources and pathways of tumor cell dissemination and initiation of LRs, pLNs, METs. (B) Tumor size dependent MET and pLN initiation and survival. I: METs can be initiated until the removal of a PT. II: PTs will initiate up to pT1c about 10% METs, 1.5% are already T-N-M1. III: During growth up to pT2, 4.5% become T-N-M1, 25% METs and 20.3% pLNs are additionally initiated which can be avoided by early detection at pT1. Equal rapid MET growth is outlined. Because of successful ATs, the percentages depend on the reference period and can therefore vary. (C) Relative* survival depending on tumor diameter for T-N-M0 PTs. (D) Relative* survival depending on the number of pLNs^18^. The arrows are are of equal length. The arrows show the decreasing risk of the increasing number of pLNs. (E) Lymph node infiltration. Distribution of the number of pLNs in dependence on tumor diameter (n=30,170). Cumulative observed data (dotted lines) and fitted Gompertz functions (Gf) (solid lines) show for each TD the percentages with >npLNs. The dotted black line estimates the proportion of patients (0pLN*) with occult METs initiated during 0pLN-status. The length corresponds to the MET-risk of 0pLNs and 1pLN in Fig. 2D. The black line is the fitted Gf for the observed tumor specific 15-year mortality (diamonds) depending on TD. The stacked barplot visualizes the distribution of pLNs at 22.5mm TD. (F) Overall survival for patients with pT1-2 PTs, breast conserving surgery and irradiation. The blue dotted and dashed subgroups are samples with the same distribution of age, pT and LNs as the subgroup without radiation (brown curve), which is further divided according to the occurrence of LRs. (G) Relative* survival for pT1c and pT2 PTs and 4 time periods from 1978. The improvement after 15 years is about 10% absolute in both subgroups. (H) Growth trajectories for MET and 2ndPTs. Each trajectory describes the number of TCs (log scale) as a function of a median growth time (8 MET or 12 PT years) relative to the PT. The age of METs or 2ndPTs at the time of diagnosis of the 1stPT indicate the age scales. Blue trajectories represent synchronous events, red occult foci and black ones for 2nPTs, which are initiated after PT diagnosis. The age distributions of METs are snapshots at the time of PT diagnosis and a delayed time afterwards. Since no new METs are initiated, there are no small METs for delayed ATs and the larger ones (blue circle) have already been discovered. *The relative survival is an estimate for tumor-specific survival and is calculated by dividing the overall survival after diagnosis by the survival observed in the general population with comparable age distribution.

### Initiation of local recurrences

When TCs migrate but remain close to the PT, they can initiate true LRs. These usually occur within 3 centimeter (Fig.2A), the target area of the boost irradiation. The shared microenvironment of the PT and the LR forms a supporting niche providing particularly favorable growth conditions. This was shown in historical data from the NSABP-B-06 study with 39.2% LRs after breast conserving surgery without irradiation.^12^ However, the term LRs is inaccurate because LR such as METs or pLNs can also occur synchronously with PTs in about 18% and are then referred to as multifocal PTs.

In total, four types of LR can be distinguished: growing residual tumors from positive margins, true PT-near LRs, and independently initiated ipsilateral 2ndPT which, if synchronous, are also called multicentric PTs (Fig.1).^13,14^ A fourth type may be foci that emerge through self-seeding.^15,16^ Since these are initiated by TCs, which find their way back through circulation, they likely exhibit similar characteristics as true LRs but without the proximity to the PT (Fig.2A). In addition, this form of self-seeding can initiate contralateral foci. It can be assumed that tumor cell dissemination from PTs must have already been taking place for some time if LRs appear simultaneous with PTs. In parallel, as data from the MCR shows, initiation of pLNs becomes more likely with about 15% more pLN findings for pT1-2m (multiple) PTs.^17^

### Initiation of positive lymph nodes

TCs can infiltrate LNs through lymphatic dissemination. This LN infiltration is also a stochastic process over time, with the number of pLNs representing discrete successive steps.^18^ This process can be described by a simple Markov model with transition probabilities for each status with npLN to reach the (n+1)pLN status by one millimeter growth. Fig.2B shows LN infiltration depending on pT-categories. In pT2-PTs more than half (26.2%) of the 46.5% pLNs are likely to have been infiltrated by TCs during growth up to pT1c. In the subgroup of hormone receptor (HR) positive non-advanced PTs, the percentage of patients with more than npLNs can be described by Gompertz functions with 3 parameters. Cumulated and complementary to 100%, Fig. 2E shows observed and fitted data for 0 to > 10pLN. For 0pLN results: y (≥0pLN,%) = 100 – 75.4*exp(−2.82*exp(−0.063*TD) (TD: 2 – 90mm).

In addition, large PTs can develop the required driver mutations at a later point in time but the infiltration process is not significantly affected. As the TD increases, the fraction with >10 pLNs increases at the expense of 0pLNs, the proportion of which varies between 0.05% and 1.75% per millimeter. However, the proportion with 1, 2 or 3pLN is largely independent of the TD for larger PTs. In the 0pLN status, some of the PTs have begun to disseminate MET-competent TCs (Fig.2D). In PTs with negative sentinel LNs approximately 15.9% of isolated TC or micro-MET are detected, the proportion of which is shown as a function of diameter in Fig. 2E as the 0pN* line and corresponds to the proportion of 1pLN.^19^ The sum of all pLNs that initiate growing and continuously disseminating PTs in 100 patients can also be described with a Gompertz function: sum (pLNs /TD) = 646 * exp (- 3.91 * exp (−0.041 * TD)) (2 <TD <90mm).^18^ The asymptote of 646 pLNs is a biological characteristic, resulting from the proportion of the subgroups 0pLN up to >10pLN in large tumors. Because of the maximum of about 30 LNs the mean value for >10pLNs is 17, and is largely independent of the TD.

### Initiation of metastases

When are METs initiated? As described above, clinical findings on the effect of pT-category show that for pT1-PTs about 13% metastasize despite ATs and the tumor-related deaths occurs in the first 15 years (Fig.2G). When comparing such results, the time reference of the cohort and therefore the ATs of the 2000s should be taken into account because progress has been made with successful innovations. If PTs are diagnosed later as pT2-PTs with a mean TD of 28mm, the METs already initiated up to a tumor size of pT1c continued to grow and 25% new METs not eradicable by ATs are initiated after pT1c size (Fig.2B).^20^ In the pT2 category the proportion of advanced PTs has already reached 4.2% and the MET-free interval shortens to 4.2 years. The time scale of the MET-P in pT2-PTs illustrates that advancing the PT diagnosis avoids METs and results in a longer MET-free interval of 5.1 years. This constitutes a lead time effect for the subgroup with unavoidable METs, which are initiated up to a tumor size of pT1.

The initiation of MET is similar to that of LNs. At some point a TC succeeds in infiltrating a distant organ site, perhaps cooperatively as a homogeneous cluster or with other heterogeneous circulating TCs (polyclonal origin).^21^ Therefore, METs are already initiated in the pN0* phase as shown by the doubling of mortality from 0 to 1pLN(Fig.2E). Thereafter, additional METs can be initiated sequentially by a PT, which can acquire further mutations in the meantime (Fig.1). Therefore, multiple METs in one or more organs and also in different pLNs can be genetically different.^3,22^ In addition, different areas of a PT can already be assigned as source of TCs.^23^

### Combinations of initiated sMETs

If a PT disseminates MET-competent TCs in principle all three sMETs can be initiated. LR are more common than pLNs, which in turn are more common than METs because TCs have to overcome additional lymphatic and hematogenous barriers, extravasation and colonization into organs.^1^ All combinations of sMETs in any order are possible. In the absence of a variant, not only chance but also necessity may play a role, which may be related to MET organ-specific properties of TCs. ^24-26^ The competing initiation is illustrated in the 15-year relative survival which is 88.1/93.8/87.9/84.7% for pT1c and all/pN0/1pLN/2pLNs respectively. The corresponding values for pT2 are 67.4/80.7/72.9 /64.6% and shows the high proportion of pLNs and METs initiated early and growing in parallel with the PT.

### Initiation of primary tumors

The age-dependent incidence also reveals the initiation of 1stPTs^20,27^, that occurs years before according to the growth duration of the PTs. About 2% of patients have bilateral PTs which must be initiated nearly at the same time and then grew in parallel. The incidence of contralateral 2ndPT and PTs in prevention studies^28^ show that year after year these PTs are diagnosed and must have been prevalent in the patient cohort according to the duration of the growth of PTs (Fig.2H). In high-risk groups^29^ with BRCA mutations the incidence increases to 60-80% compared to the lifetime incidence of approximately 12.4% in the normal population.^27^ In particular, BC patients have a 3-5 fold risk of developing 2ndPTs compared to the incidence of 1stPT in the normal population.^30^

### Source and time of late MET initiations

#### Dormant TCs

DTCs could be an inexhaustible source to initiate MET after PT removal.^31-33^ But the biology of these DTC and their natural history over a patient’s lifetime is largely unclear.^34,35^ The only argument for their existence is the detection of vital TCs in organs.^36^ Such TCs exist and are a prognostic factor but probably not a relevant cause of METs. Improved diagnostics reveal more synchronous LR and advanced PTs. More elaborate preparation of sentinel LNs detects isolated tumor cells. Occult METs of all sizes are also present in organs as the distributions of MET-free survival show (Fig.3B). Late METs are initiated shortly before PT diagnosis and their MET-free periods of 10 or more years need not be bridged with dormancy. Successful ATs have been shortened to few months from years ago.^37^ This implies that even in large studies no relevant risk by METs could be demonstrated that would have been initiated by dTCs in the previously longer treatment phase. Therefore, also an ectopic evolution of TCs in niches should not be a relevant step in the MET-P.^6,38^ Thus, only the risk of MET initiation by TCs of the three sMETs has to be considered (Fig.2A).

**Figure 3:**
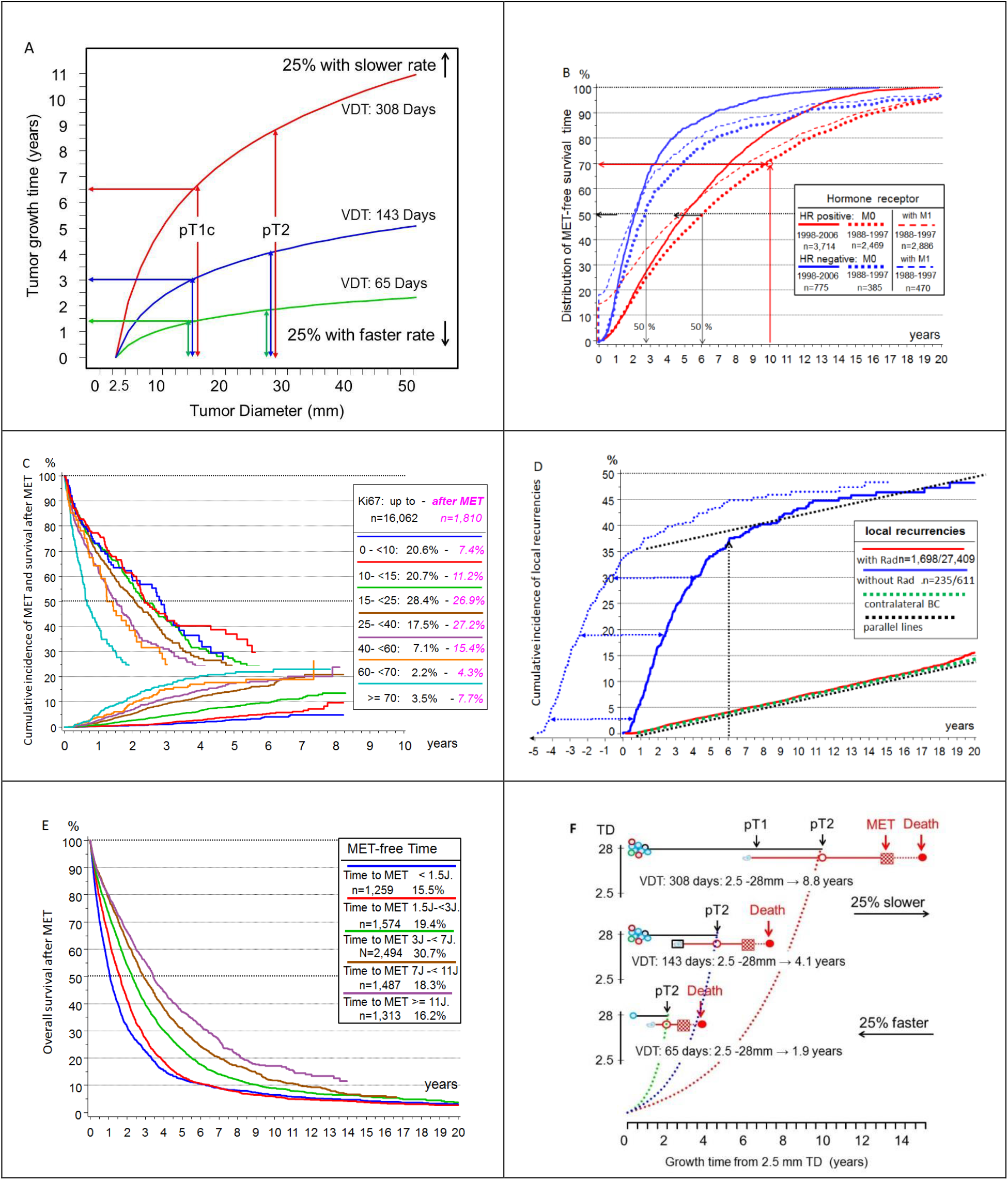
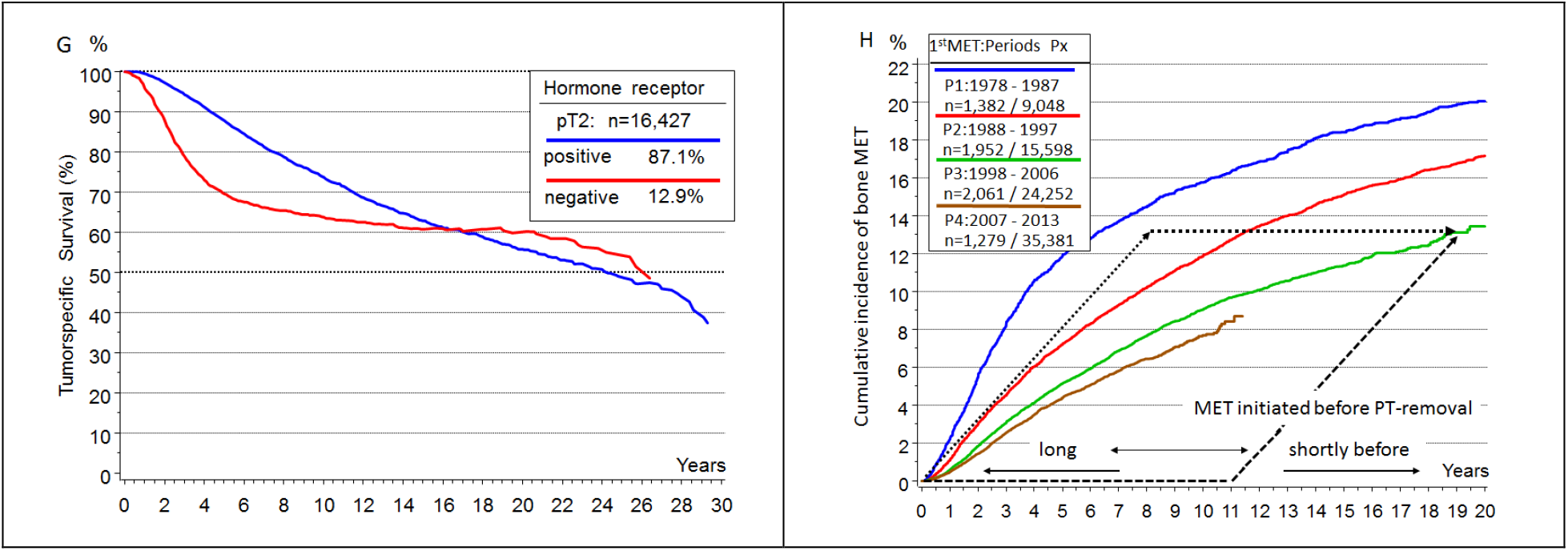
3A – 3H combined **(A)** Volume doubling and tumor growth. Growth durations of PTs from 2.5mm depending on estimates of the 25% / 50% / 75% percentiles VDTs according to Weedon-Fekjaer^55^. The variability is illustrated by the growth times from 2.5 mm up to pT1c or pT2-PT. **(B)** Hormone receptor dependent MET-free survival. Distribution functions of the MET-free times for T-N-M0 and HR+ (red) and HR-(blue) PTs from the time periods 1988-97 and 1998-2007. Since METs also occur after 20 years, the distribution reaches 100% after the maximum follow-up time, e.g., after 20 years. About 30% of the MET appear after 10 years. The dashed distributions also show the proportion of primary MET.for 1988-97 cohorts. **(C)** Cumulative incidence of METs and survival after MET depending on KI67 (available since 2012). The median survival at T-N-M0 in the event of tumor-related death for the two outer subgroups is approximately 3.1 and 1.5 years. **(D)** Cumulative incidence of LRs after breast conserving surgery with and without irradiation. Black dotted lines are parallels and show that late LRs increases in both groups comparable to the incidence of contralateral 2ndPT. The dotted blue curve is similar to those for LRs without irradiation and is the fictive initiation of true LR after forwarding the twofold LR-free time of about 26 months. **(E)** Overall survival from MET of HR+ PTs as a function of MET-free time. PTs are diagnosed between 1988-2006 and n=8,127 METs evaluated. The differences of PTs of the 1st and 5th Quintile (<1.5, ≥11 ys) are 12.4/36.9% for 5 years survival, 23.9%/45.7% for pT1 and 18.3 / 42.4% for pN0. **(F)** PT/MET growth relation principle. Relation of tumor growth of PTs and METs (pT1,pT2), MET-free survival and post MET-survival (red) at different growth rates.^49^ **(G)** Relative survival of HR+ and HR-PTs. **(H)** Time trend of bone MET-free survival. Cumulative incidence of bone METs as a function of 4 time periods since 1978. (Kaplan-Meier method without concurrent risks) The black lines sketch fictitious selective eradication of early (dashed) or late (dottet) initiated METs alternative to the green line.

### True local recurrences

True LRs are not considered a source of MET because 39.2% LR after breast conserving surgery did not result in a higher mortality in the seminal NSABP-B-06 study.^12^ The high 4:1 risk found in meta-analyzes has meanwhile been reduced to the subset of pN+ findings.^39,40^ Such a constraint is not plausible because survival curves show a continuously increasing MET-risk that starts at 0pLN* (Fig.2E). This can be also shown by data of the MCR: in the subgroup of non-irradiated patients where 38.7% of LRs occur without any effect on survival, which reproduces the results of the NSABP-B-06 study.

### Positive lymph nodes

Synchronous pLNs are not involved in MET seeding^41^, nor has any robust data for cascading initiation been presented to date.^42,43^ Long-growing PTs can infiltrate 10 or more LNs. If after a sentinel extirpation a LN recurrence is diagnosed it is often a singular pLN, which does not infiltrate the subsequent LN network like PTs. As mentioned above, METs are also initiated when no LNs are involved (Fig.2D). In none of any solid tumor radical LN dissection has resulted in a survival benefit. This has been demonstrated for BC and more than 10 other tumor types and can also be logically deduced from Fig.2D.^18,44,45^ The more than a century old hypothesis of cascade-like spread cannot be sustained.^46,47^ Survival curves in Fig.2D-E show that the additional MET-risk decreases with each additional pLN. Therefore, pLNs are dead ends within the MET process even though animal experiments suggest a cascading spread is possible.^4,48^

### Metastases

METs disseminate TCs with prognostic and predictive relevance.^49,50^ However, cascade-like initiation is clinically difficult to detect because, due to growth periods, patients would generally not experience METs initiated by METs. However, there are studies which claim that each sMET can initiate tertiary METs. Estimates of initiation and growth of METs are usually not considered. In addition, any study must concede that “we cannot formally exclude an alternative explanation for the observed patterns, that each of these METs has seeded from an undetected subclone in the PT”.^51^ This risk can also be largely ruled out because multiple MET in different organs and their segments are often genetically different.^23,52^ MET surgery data also points against this argument because, after R0-MET resections, no new proximal METs limit the successful local resection as in 60% LR (inclusive multifocal LR) PTs without irradiation.^53^ Taken together, this data supports the hypothesis that LRs, pLNs, and METs themselves most likely cannot initiate clinically relevant new METs. Therefore, all sMETs and also 2ndPTs of at least the next 10 years are already prevalent at the time of PT diagnosis (Fig.1), which are targeted with varying degrees of success by primary systemic ATs and/or radiotherapy.

### Growth duration

The mechanisms of tumor growth remain largely unknown. From the smallest clusters of cells to angiogenesis in the more advanced disease phase and thereafter, tumor growth varies due to differential cell divisions inherent to the molecular subtypes and unknown apoptotic rates. In addition, there are dependencies for extravasation and colonization on the tumor microenvironment^26^. Nevertheless, growth duration can be estimated based on the initiation period (Fig.2B), prevalence, PT- and MET-free durations, and can be transformed in volume doubling times (VDT). The growth of tumor foci can be described by growth trajectories^54^ that show the increase in the cell number over time (Fig.2H). The growth will be mostly exponential. With the logarithm of the number of TCs, a straight line is obtained for which an asymptomatic logistic growth as in Fig.2H seems plausible at the beginning and at the end. For a cohort, the age distribution of occult METs at the time of diagnosis and their occurrence during the course of the disease can be elucidated.

### The growth of PTs

In parallel with PTs, the METs already initiated are also growing. If there are about 1.3% M1 in pT1a but already 12.3% for pT3 and varying MET-free intervals depending on TD, then the growth of the PT is also a chronometer for simultaneously growing MET (Fig.2B). There are three approaches to this process: Estimates from screening data showed a median VDT for women aged 60-69 years between 10 to 20mm within 143 days.^55^ If these growth rates also apply starting from a diameter of 2.5 mm (pT1a) the variability of the VDT with 25%/50%/75% percentiles of 65/143/308 days results in a growth time between 2.5 to 15 mm (pT1c) in 1.4/3.0/6.5 years (Fig.3A). The 25% percentile at 1.4 years is consistent with approximately 25% that occur as interval cases in a biennial screening.^56^ The growth time of HR+ occult BCs, which can be influenced by hormone replacement therapy, are correspondingly longer. The mean age at diagnosis 60.9/61.7/64.6 for pT1a-b/pT1c/pT2 also largely reflects PT growth. The great variability of VDT is also apparent for molecular subtyping.^57^ Since sMETs are rare in pT1a-PTs, it follows that all sMETs usually grow faster than PTs.

The second approach is provided by prevention studies. With the above VDTs of 65/143/308 days and 32 VD, a pT1c-PT would grow from TC to diagnosis 5.7/12.5/27.0 years. In keeping with this, prevention studies show a continued reduction in incidence even 15 years after the end of a 5 year endocrine chemoprevention.^28,58^ Fifteen years of PT growth corresponds to 171 days of VDs for 32 cell divisions. If the above 143 days are assumed this would result in a median growth of 12.5 years. That means that at PT diagnosis most of contralateral 2ndPTs for the next 12.5 years are already prevalent. In about 2% of cases synchronous PTs are detected. At the beginning of the mammography screening recommended for women aged 50 and over, 4% PTs are not yet detectable but already prevalent, and will occur in the next 12.5 years. The short-term increase of incidence by postmenopausal estrogen plus progestin therapy and its decline after weaning can also be explained by the prevalence and growth stimulation of PTs.^59-61^ The third approach is provided by the estimation of MET growth from MET initiation to PT diagnosis.

### The growth of METs

The growth of METs can be estimated if the 4 reference points are observed (Fig.2B): a lower limit for initiation starting at about 1mm tumor size, the timing of PT and MET diagnoses and the tumor-related death. The growth time of METs consists of the growth up to PT diagnosis and the MET-free time afterwards. In the special case of primary MET the MET has grown parallel to the PT (Fig.2H). If METs were initiated at 2.5mm, they would grow a median of 3.0/4.1 years parallel to pT1c/pT2-PTs and therefore must have more genomic differences to the PT than late-initiated METs.^62^ In pT2/pT3/pT4-PTs about 4/12/27% are primarily metastasized. In the case of T-N-M0-PTs the distribution function of the MET-free time with a follow-up of more than 20 years results in about 30% METs >10 years and a median of 6 years (Fig.3B).^63^

If T-N-M1 cases are also included, the distribution begins with a step corresponding to the proportion of M1 in all courses of disease with MET (Fig.3B). A significant regression results with the Gompertz function y(%) = 98.9*exp(−1.82*exp(−0.19*t)) (−10 - t(years) – 20), MET with HR+ PTs, 1988-97). The median MET-free times with T-N-M1 result in 2.0/4.8 years for HR neg/pos PTs or together 4.4 years. The double is 8.8 years or 100 days for 1 VD and a plausible estimate for a median MET growth. The MET growth varies similar to PT growth. The MET-free time and the time after MET already differ by a factor of more than 2.4 solely based on the PT HR status (Fig.3B-C). But there is also a great variability within these subgroups. If the MET-free time is divided into quintiles, then the 5-year survival after MET is 10%/40% for the outer 20% limit values of approx. <1.5 and >11 years (Fig.3D).^64^ The longer the MET-free time the more favorable the prognostic factors of the PTs are.

This contradicts a continuous MET initiation from dTCs after R0 removal. METs that would not be initiated until 5 or 10 years after PT diagnosis would have to grow very fast and most of them would have to originate from triple negative PTs, which is not observed. Growth rate variability reflects the distribution of event-free times, which are positively skewed with a long upper tail for METs that occur without dormancy phases after 15 years or more. Despite the increase in MET frequency with TD, MET is an autonomous process within molecular subgroups. Larger PTs do not initiate more aggressive METs. According to this principle METs initiated early or later on grow comparably fast (Figs.2H, 3C), therefore MET-competence of TCs is a qualitative not a quantitative characteristic. METs can acquire additional mutations^3^ (Fig.1 and become a moving target for precision oncology, which does not seem to affect growth rates.

### The growth of LRs

The growth of LRs can be read from two time durations: Late initiations of true LR occur immediately before PT removal and are likely to have the longest LR-free times. This growth period of true LRs can be read from the breast conserving surgery studies with and without radiation^39^ and is about 6 years (Fig.3E) because thereafter the slope of the cumulative incidence of LRs is the same in both groups. The earliest initiations of true LRs results in 18% multifocal findings and without them the median LR-free time is approximately 2.5 years and independent of follow-up (Fig.3E), while median ipsilateral 2ndPT-free time increases to half the follow-up time due to continuous initiation of PTs (Fig.2H).^65,66^ Six years of growth results in a VDT of about 68 days and 32 VDs, almost 2.1 as fast as that of the PTs.

### The growth of pLNs

The growth of pLNs cannot be easily estimated because of the lack of robust data on pLN-free time due to successful ATs and LN dissections. According to data of the MCR in 100 patients with pT1b-BC (7.5 mm), 13% pN+ and a total of about 36 pLNs are to be expected. If PTs reach a diameter of 15mm after about 14 months, 42 more LNs are infiltrated, in total 25% pLNs. In today’s serial sentinel preparation, small PTs have about 12% macro-METs and almost as many isolated TCs and micro-METs.^67,68^ If the 12% newly infiltrated LNs had a size of 0.2mm at 7.5mm (border between ITC and micro-MET) and then grew to a size of 2mm, this results in a VDT of 43 days, 3.3 times that of PTs.^43^

### MET-free and post-MET survival

The survival of a MET-related death is composed of three periods of MET growth (Fig. 2B), the time to PT diagnosis, the MET-free time, and the post-MET survival. If PTs from 1mm TD can initiate MET, then this will become even more likely with an increasing size of the PT. The longer a PT disseminates, i.e. the larger the PT, the longer the occult MET growth and the shorter the MET-free survival time. This also applies to pLNs. Of all METs 35% occur at 0pLNs. The MET-risk increases when micro-METs are already detectable in LNs and it doubles when the first LN is positive (Fig. 2D).^43,67,68^ If more LNs become positive these occult METs continue to grow, the MET-free intervals become increasingly shorter. In 0/2/8-10 pLNs, primary METs are diagnosed in 0.7/4.1/14.2%.The number of pLNs is the most important clinical prognostic factor, it is a chronometer for MET but not its cause.^43,44^ Therefore, evidence for extensive dissection for MET risk reduction is lacking in the most common solid tumors, although LN extirpations may be important for regional control.

True LRs are caused by disseminating PTs, which can also initiate METs in parallel to LNs and organs. As in the NSABP-B-06 trial there is no MET-risk in MCR data. Because of the parallel MET, the group with rad has a less favorable prognosis (Fig. 2F). In addition, the growth times of the MET do not fit the published 4:1 survival effect^39^ that should occur without irradiation due to the not avoided LRs and their caused METs. The lack of MET-risk is also supported for synchronous true LR, i.e. multifocal findings, for which no increased MET-risk could be detected.^69,70^ Progression data in Fig.3B-D are easier to interpret if the progression-free times are not considered from diagnosis but from the initiation of the progressions, which was about twice the median MET free time before the respective MET discovery (Fig.3D). It is important to note the variability of survival time after MET and the correlation of time to MET because MET discovery does not alter MET growth. Post-MET survival reveals the influence of the various growth factors. Median post-MET survival varies e.g. at Ki67 between 0.7-3 years (Fig.3C). Since growth also varies by a factor of 10 or more with a qualitative characteristic such as HR status, there must be a quantitative molecular biomarker for the growth rate that correlates higher than the combination HR and Ki67.

Conversely, the duration of the MET free period is also another prognostic factor for survival after MET (Fig.3E). However, this correlation is also dependent on the extent of the MET and its localization. For example, the median survival after bone and CNS-MET differ by the factor 3.^71^

### Hypotheses about tumor growth and the MET process

Available evidence from well-known data about PT findings, age-related incidence and the course of disease provide a plethora of facts that can be derived from five hypotheses on the initiation and growth of PTs and METs and the effect of today’s successful neo- and adjuvant therapies. Neither initiation of METs by secondary foci nor a long-term delay through dormancy could be convincingly shown so far. The prevalence of METs at PT-diagnosis could be summarized in an (1) ***pre-diagnosis MET initiation principle of the primary tumor***. Growing PTs continuously initiate life-threatening METs. Derived from tumor-specific mortality, the cumulative incidence of MET as a function of TD is nonlinear and follows a Gompertz function. This is a (2) ***growth dependent metastasis principle***: With every millimeter of a growing PT the proportion of METs increases, with early and late MET being less frequent compared to the maximum at the inflection point of the Gompertz function, with today’s data at 21mm. This random process leads to a variability in the age of the MET at diagnosis and the MET-free time, which together give a mean duration for MET growth of about 8.8 years (Fig.2H).

Growth rates of PTs and their METs can each differ by a factor of 10 and more with a long upper tail (Fig.3A). Despite the great variability and molecular heterogeneity, the dissemination of TCs and initiation of pLNs and METs, depending on TD, are likely comparable. The processes only run at different speeds, which is a (3) **time lapse or time dilation principle**. Therefore, the properties of 25% interval PTs are comparable to populations without biennial screening despite their rapid growth^56^, or early advanced PTs are not predominantly triple negative PTs.

Dissemination, acquisition of MET-competence, MET initiation, and growth rates are associated with prognostic factors. HR or HER2 status define homogeneous subgroups in which the principles apply and are the first targets for personalized therapies. The existence of homogeneous, clinically relevant and yet to be discovered subgroups is the basis of progress, (4) a **subtype existence principle**.

Triple negative or luminal A-PTs usually do not initiate contrary METs. That is, the relationship of the growth rates of PTs and their METs varies only slightly also between the subtypes (Fig.3F). The relationship must be greater than 1 because there are M1-PTs and it cannot be 10 or more because then there would be mostly M1-PTs. The estimate of about 8.8 years of growth for MET and 12.5 years for PT results in the factor 1.4, which, however, depends on the length of the follow-up. This growth relation can be seen as a (5) ***robust PT/MET growth relation principle*** and independent of the subtype. The tumor-specific long-term survival supports these principles and they are a challenge for gene expression tests (Fig.3G).^72^

### Effect of adjuvant and neoadjuvant treatments

The principles of MET-P explain successes, limitations, and opportunities of ATs and, conversely, successful ATs confirm the principles. All secondary foci are already prevalent at primary therapy because LRs, pLNs, and METs cannot be initiated after removal of the 1stPT. This leads to three questions for successful ATs: 1. From and up to what size are METs eradicable? 2. How many METs can be eradicated? 3. How long does treatment have to be given? The same questions also apply to occult contralateral 2ndPTs.

### Eradication of primary tumors

Complete pathological remissions of over 50% can be achieved with neo-ATs in a few weeks.^73-75^ Endocrine therapies achieve a less complete remission, but downsizing and prolonged localized control of the PT are possible.^76-79^ The lower efficacy of endocrine neo-ATs depends on the size of PTs, because the reduction of the incidence by endocrine chemoprevention starts after a short delay.^28,80^ The delay is less than 3 VDs and can be interpreted as lead time effect via the inclusion criteria of a negative mammography exam. Thus, prevention eradicates PTs quickly, especially under aromatase inhibitor.^81^ Since further 1stPTs are initiated during the 5 years in the prevention studies, the eradication of these smallest clusters becomes apparent 15 years later. This means that the prevention successfully eradicates about 50% PTs of all sizes up to a threshold of almost detectable 1stPTs. This is exactly how endocrine ATs act on 2ndPTs, and this has been confirmed many times on the contralateral 2ndPTs.^82^ PTs diagnosed according to the median growth times of PTs in the next 12.5 years are already prevalent at the start of chemoprevention. It follows that there is no rationale for 5 years of preventive therapy. The facts suggest chemoprevention of a few months.

### Eradication of metastases

Overt METs have not been eradicable until today. This also applies to nearly detectable MET, because in clinical trials with (neo-) ATs the survival curves of MET-free time do not separate in the first months.^83-85^ Thus, there is a size-dependent upper threshold for effectiveness comparable to chemoprevention. A lower limit is not recognizable from our historical data because successful ATs have achieved a uniform eradication of even the smallest METs (Fig.3H).

This also follows from the distribution of the number of pLNs: the fraction with 0pLN* and 1-3 pLNs is largely independent of the TD (Fig. 2E), but the mortality increases with each additional pLN (Fig.2D). In these subgroups, there are a similar number of small METs which can be destroyed by ATs. This explains the more than 10% improvement in survival seen in the past decades, regardless of the PT size.^64,86^ With a favorable prognosis (0pLN, pT1), the relative 5-year survival today reaches almost 99% (Fig:2C-D-G).^27^

Alternatives such as the selective eradication of early initiated METs with few mutations or late-onset MET due to smaller foci are not recognizable (Fig.3H). However, the already mentioned successful ATs of the past few decades provide a selective eradication of METs: Nowadays, METs in bone or lung are about 50% or 30% less frequent, whereas there is no evidence of clinically relevant eradication in the liver and CNS METs until now. Such successful ATs produce a paradoxical effect. If the early bone METs become less frequent and the progression begins with the later CNS METs, then MET-free survival becomes longer and post-MET survival becomes shorter.^64,87^ In contrast, the MET pattern in T-N-M1 (at diagnosis) has not changed in the last decades because it is untreated and mirrors the biology of BC.^71^ A rationale for the duration of ATs is not known.

The process becomes more understandable when the effect of delayed-onset ATs is considered.^88^ If surgery is postponed by a few weeks, further METs and pLNs are initiated with each additional millimeter of TD. This is the contrary of the logic of screening to shorten the duration of the dissemination of the PTs and thereby prevent METs (Fig.2B-C). If ATs are started with a delay, there is initially the same effect as if they were started immediately.^89^ However, because all prevalent METs continued to grow during the delay, there are fewer eradicable METs later on and some have exceeded the eradication threshold (Fig.2H).^89,90^ That also suggests the effectiveness of short treatment durations and justifies the question about ATs: “How short is short enough?”.

### Eradication of LR and pLNs

*The eradication of occult LRs by irradiation is excellent*.. Radiation therapies even made breast conserving surgery possible due to its efficacy in faster growing LRs compared to PTs which are also prevalent in the form of ipsilateral multicentric 2ndPTs (Fig.3E).^12^ Therefore, ATs may improve local control of multifocal and multicenter LRs, the different origins of which should be shown by their clonality.^91^

*Regional control of pLNs* is also achieved with today’s surgical and neo-ATs making a LN relapse in <5% of cases during a time span of 15 years a rare event. Survival is optimal if no residual tumor remains after neo-AT primary and in the LNs.^73,75^ Thus, ATs also contribute to regional control because in the absence of an axillary dissection they act as neo-ATs on not removed LNs with isolated TCs and micro-METs.^44,92^

### Hypotheses about neo- and adjuvant treatments

Treatment cohorts of MCR and clinical studies also suggest five principles: **(1)** Endocrine chemopreventions can eradicate about 50% of 1stPT and 2ndPTs, regardless of whether the PT develops during the therapy, is already prevalent and very small or almost detectable. **(2)** 50% complete pathological remissions are possible with neo-ATs. **(3)** Neo- and adjuvant therapies can eradicate organ-specific METs of all sizes up to detection limit. **(4)** The eradication of PTs and METs is achieved quickly as shown by the opening incidence and survival curves and implies the challenge for the treatment duration.**(5)** The rapid eradications make intrinsic resistance in PTs and METs more likely than a therapy-induced resistance.^93^

## Discussion

These principles of today’s treatments are evident. However, they also illustrate the limits of ATs and the necessary further development of prognostic and predictive factors. This should motivate innovative treatments. The duration of successful treatments is one important point. If there is no rationale for the duration of a treatment regimen, even with randomized trials then misuse is hardly avoidable. In particular, overtreatment is likely, but undertreatment is also conceivable.^94^ Optimizations in the sense of deescalation “how short is short enough”^95,96^ are ethically problematic, scientifically unattractive, and usually economically disadvantageous. But there are such studies that have achieved equal efficacy with shorter treatment durations.^97,98^ Real world data may help to examine possible improvements, for example, in the case of good results despite deviations from guidelines. Other examples include the use of a recurrence score and waiving of ATs in populations or the evaluation of alternative decision rules when a recurrence score is retrospectively obtained in the knowledge of follow-up.^99-101^ If growth of PTs varies by a factor of 10 or more, new biomarkers for the duration of therapies are conceivable as a further step towards personalized medicine and could be tested with registry data.^102^

It is particularly difficult to change knowledge or treatments acquired with studies. An example is the hormone replacement therapy mentioned above. There is no evidence that in 5 or 10 years HR+ PTs can be initiated and grow to detection. Prevalent PTs only grow faster and cause an increase in incidence. The risk of new PTs can only be detected in the long follow-up. Another example is extended endocrine AT^103-105^, which also contradicts above principles. Again, the studies provide correct results, but they are not interpreted correctly. Endocrine therapy eradicates occult METs on the one hand and prevalent PTs and their future METs on the other. With one short AT and intermittent short preventive therapies needed because of new PT arising throughout life, the treatment time could be reduced to at least 70%. This also reduces side effects and their complex management^106^ with the same results.Further conclusions from the principles arise for LN dissections and after-care. Despite many studies, pLNs are not a cause of MET and therefore extended LN dissections cannot be justified and the efficiency of expensive diagnostics in aftercare has not been shown.

A second point arises from the question, of what properties PTs or METs have that are eradicated or resistant. In the case of organ-specific efficacy, success is primarily linked to the microenvironment. CNS and liver are inaccessible and pharmacokinetic causes and molecular mechanisms are discussed.^107^ Early HR+, contralateral 2ndPTs that arise despite endocrine AT of the 1stPT would have to show differences from HR+ 1stPTs in the genomic landscape. This question should also be asked about METs that can only be partially eradicated. The rapid eradication of PTs^73,85^ and METs^108^ within 1-2 VDs suggests that a non-responsive tumor is more of a consequence of intrinsic resistance mechanisms than of acquired ones.^109,110^ This is also supported by the fact that there are many gene expression signatures that are predictive of poor outcome and whose genes influence estrogen receptors, apoptosis, or MET.^111^ But it is also true that many new treatments starting with MET have been tested and yield manageable benefit.^112^ If only progression-free survival and not overall survival is improved, the arduous testing as a useful AT is likely to be omitted. Meticulous observations, laboratory and mathematical modeling can help clarify hypotheses in advance and can perhaps accelerate innovations. This is supported by the hope that complexity is a manifestation of only a few fundamental principles.^2^

## Conclusions

Clinical trials and real-world data with lifelong follow-up and progression-free and post-progression survival elucidate dissemination of TCs, initiation and growth of sMETs as well as PTs. Few principles describe tumor growth and the MET-P and form a framework for scientific terms, molecular hypotheses, and treatments. Dormancy and stepwise MET initiations by sMETs are not supported by data. Also, principles about neo- and ATs can be derived from data about successes and limits of preventive, diagnostic and therapeutic interventions. Tumor growth and successful changed MET patterns question the duration of preventive and adjuvant therapies.

Primarily, real world cancer registry data should clarify how findings, treatments, or MET pattern change and which medical advances have a relevant impact on a population. With quantitative models^113^ and comparative effectiveness analyzes^114,115^ hypotheses about tumor growth and therapies can be derived and validated from the data. Feedback from this data analyzes can support delivery of care and improve its quality, especially when many outcome parameters are compared. Impulses for health care-relevant translational research can be given. Linking cancer registry data to biomaterial and sequencing data could accelerate interdisciplinary knowledge acquisition. Such expansions of the systematic use of available data can become a vast source of knowledge.

## Data Availability

Part of all data of Munich Cancer Registry is publicly accessible: https://www.tumorregister-muenchen.de/en/facts/specific_analysis.php. Detailed breast cancer analyzes can be provided by emailing the corresponding author.

## Appendices

### Disclosure

The authors have declared no conflicts of interest.

## Acknowledgment

We thank the clinics, doctors and teams of the general practitioners, the departments of pathology and radiation oncology for the systematic transfer of their findings and treatments. In particular, we would like to thank the staff of the Munich Cancer Registry for their meticulous and systematic recording and maintenance of the data. The Project Group Breast Cancer of the Comprehensive Cancer Centers has regularly discussed questions and results with the MCR. Thank you very much.

## Abbreviations

AT: neo-adjuvant systemic therapy
BC: breast cancer
HR: hormone receptor
(s)MET: (secondary) metastases
MET-P: metastasization process
LR: local recurrence
(p)LN: positive lymph node
(1st/2nd)PT: (first/second) primary tumor/breast cancer
(d/c)TC: (disseminated/circulating) tumor cell
TD: tumor diameter
VD(T): volume doubling (time)

## Supplement: 16 Figures 2A-3H shown individually with a higher resolution

**Figure 2A.**
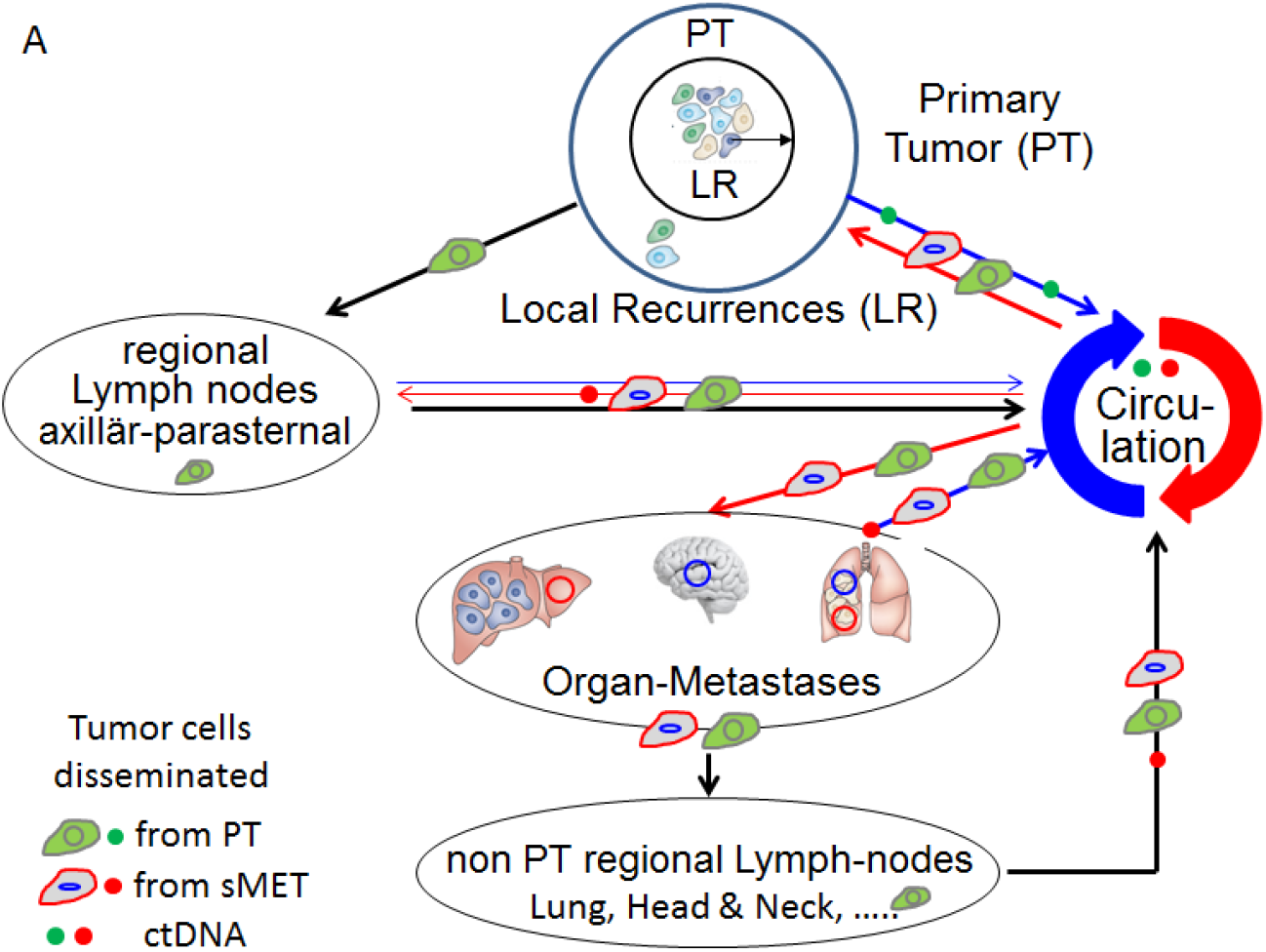
Tumor cell dissemination and initiation of LRs, pLNs, METs

**Figure 2B.**
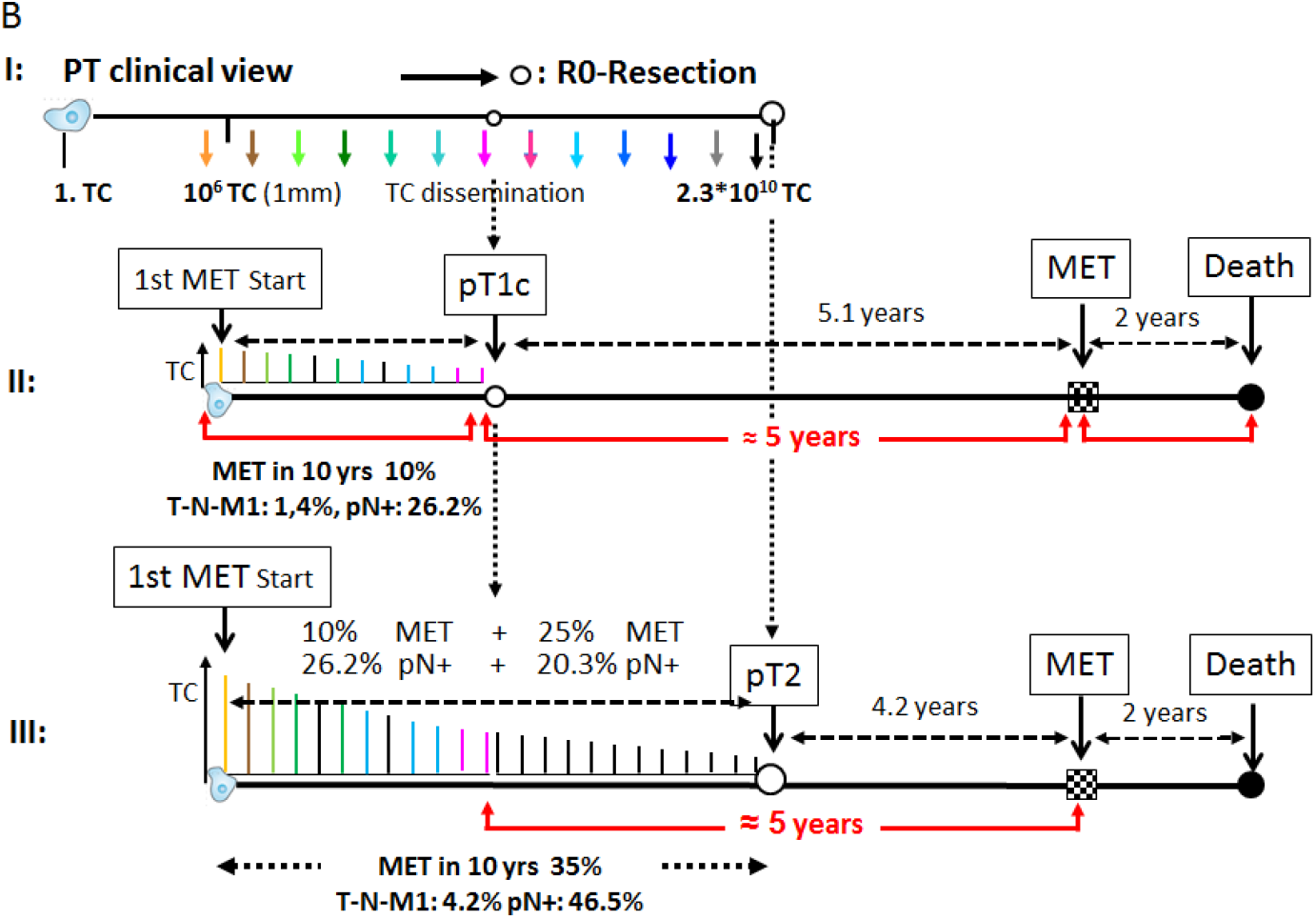
Tumor size dependent MET and pLN initiation and survival **I**: METs can be initiated until the removal of a PT. **II**: PTs will initiate up to pT1c about 10% METs, 1.5% are already T-N-M1. **III**: During growth up to pT2, 4.5% become T-N-M1, 25% METs and 20.3% pLNs are additionally initiated which can be avoided by early detection at pT1. Equal rapid MET growth is outlined. Because of successful ATs, the percentages depend on the reference period and can therefore vary.

**Figure 2C.**
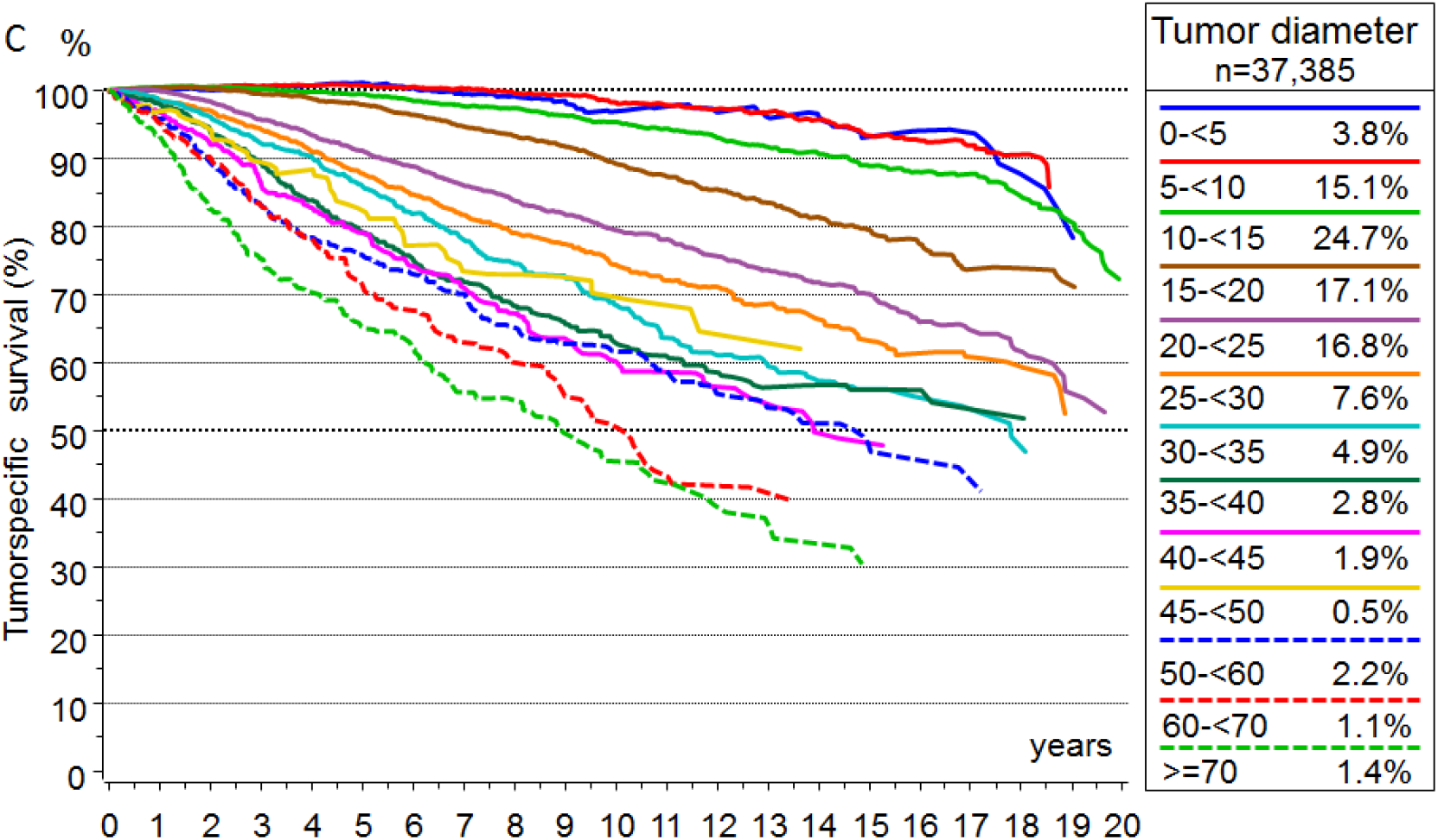
Tumor diameter Relative* survival depending on tumor diameter for T-N-M0 PTs. *The relative survival is an estimate for tumor-specific survival and is calculated by dividing the overall survival after diagnosis by the survival observed in the general population with comparable age distribution.

**Figure 2D.**
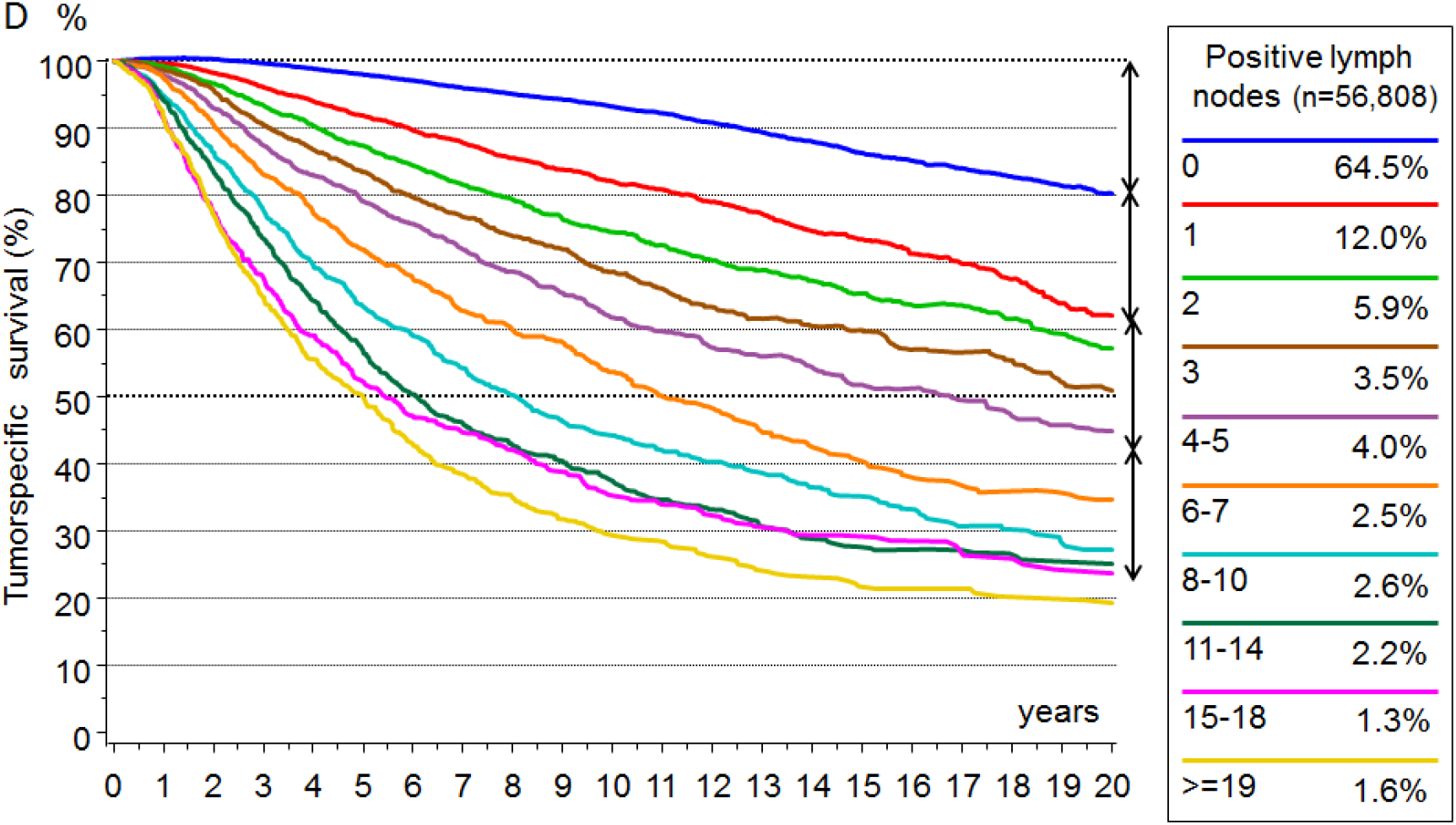
positive LNs Relative survival depending on the number of pLNs. The arrows are the same length. The length corresponds to the mortality of 20% in 0pLNs after 20 years. The arrows show the decreasing risk of the increasing number of pLNs.

**Figure 2E.**
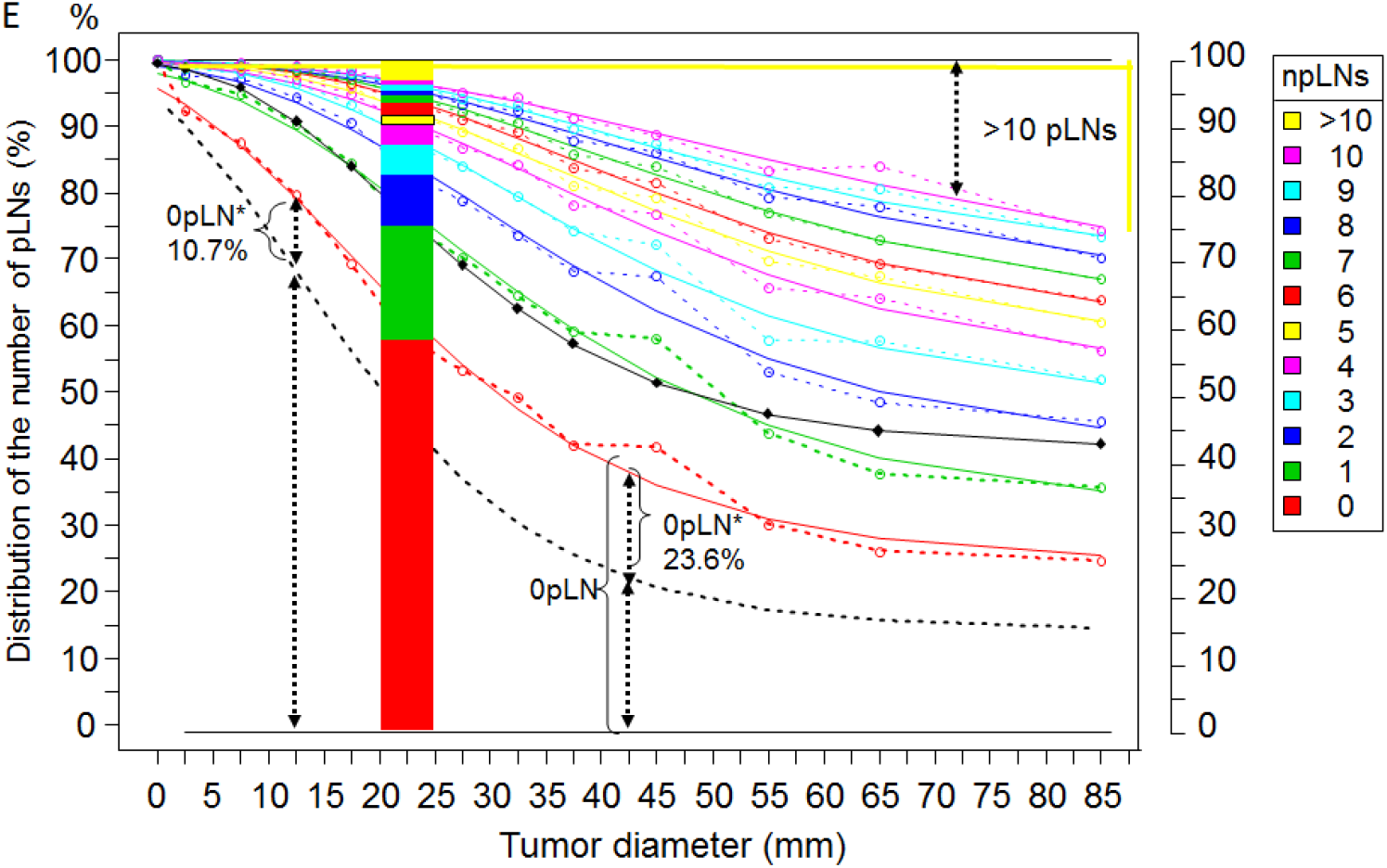
Lymph node infiltration Distribution of the number of pLNs in dependence on tumor diameter (n=30,170). Cumulative observed data (dotted lines) and fitted Gompertz functions (Gf) (solid lines) show for each TD the percentages with >npLNs. The dotted black line estimates the proportion of patients (0pLN*) with METs initiated during 0pLN-status. The length corresponds to the MET-risk of 0pLNs and 1pLN in Fig. 2D. The black line is the fitted Gf for the observed tumor specific 15-year mortality (diamonds) depending on TD. The stacked barplot visualizes the distribution of pLNs at 22.5mm TD.

**Figure 2F.**
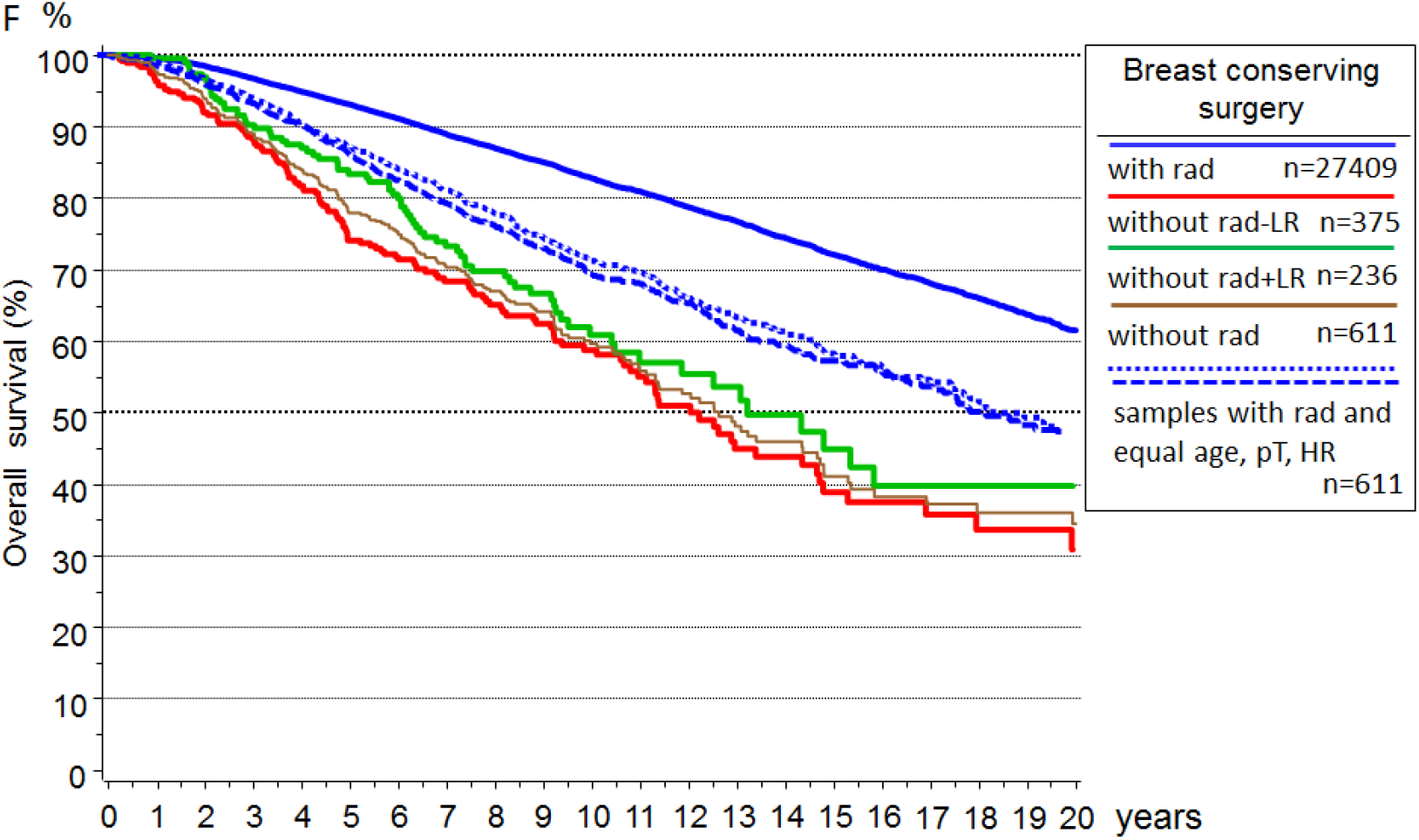
Overall survival after breast conserving surgery and irradiation Overall survival for patients with pT1-2 PTs, breast conserving surgery and irradiation. The blue dotted and dashed subgroups are samples with the same distribution of age, pT and LNs as the subgroup without radiation (brown curve), which is further divided according to the occurrence of LRs.

**Figure 2G.**
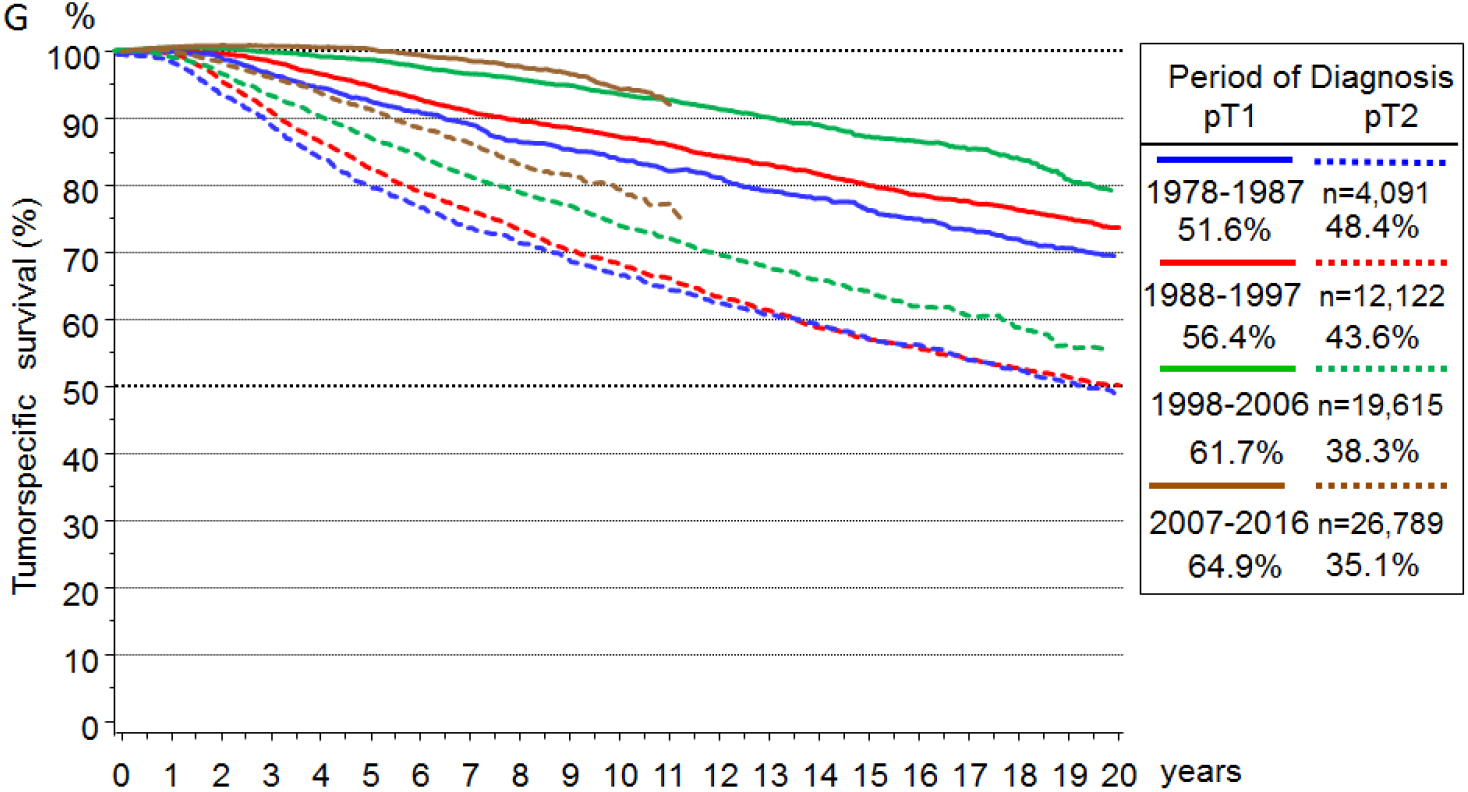
Trend: relative Survival depending on pT1-2 Relative survival for pT1c and pT2 PTs and 4 time periods from 1978. The improvement after 15 years is about 10% absolute in both subgroups

**Figure 2H.**
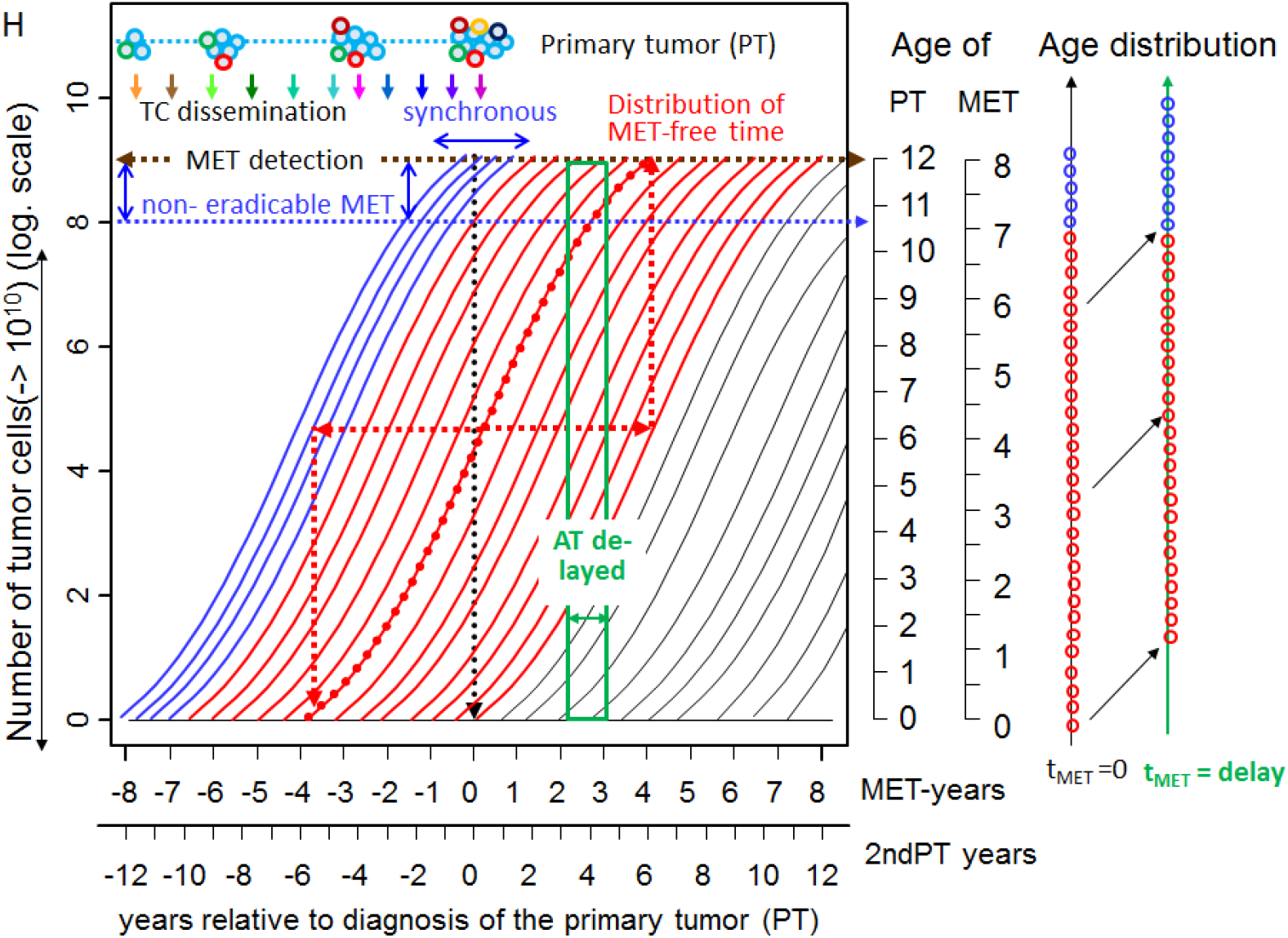
Growth trajectories Growth trajectories for MET and 2ndPTs. Each trajectory describes the number of TCs (log scale) as a function of a median growth time (8 MET or 12 PT years) relative to the PT. The age of a MET or 2ndPT at the time of diagnosis of the 1stPT indicate the age scales. The sum of age and event-free survival is on average constant. Blue trajectories represent synchronous events, black ones for 2nPTs, which are initiated after PT diagnosis. The age distributions of METs are snapshots at the time of PT diagnosis and a delayed time afterwards. Since no new METs are initiated, there are no small METs for delayed ATs and the larger ones have already been discovered.

**Figure 3A.**
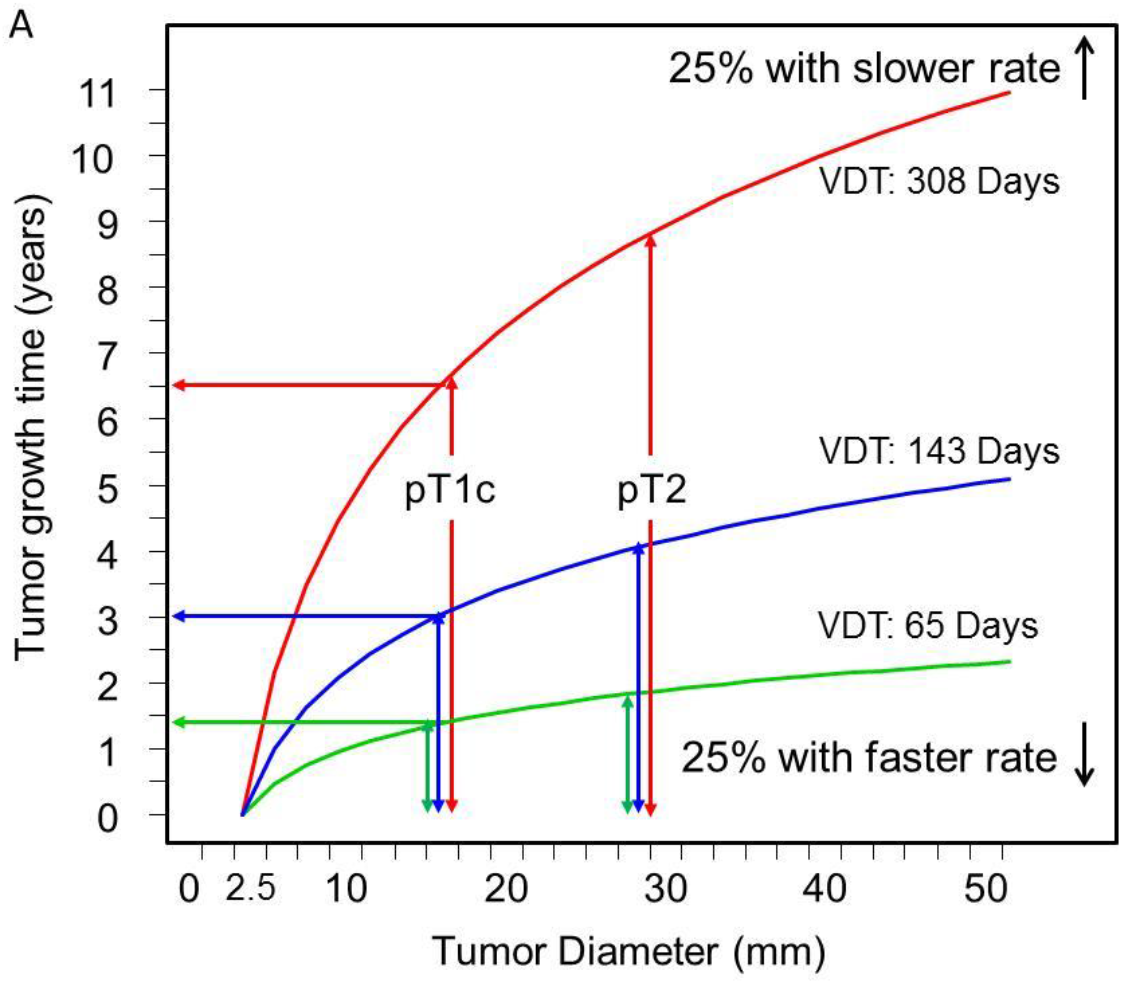
Volume doubling and tumor growth Growth durations of PTs from 2.5mm depending on estimates of the 25% / 50% / 75% percentiles VDTs according to H. Weedon-Fekjaer ^55^ The variability is illustrated by the growth times from 2.5 mm up to pT1c or pT2-PT.

**Figure 3B.**
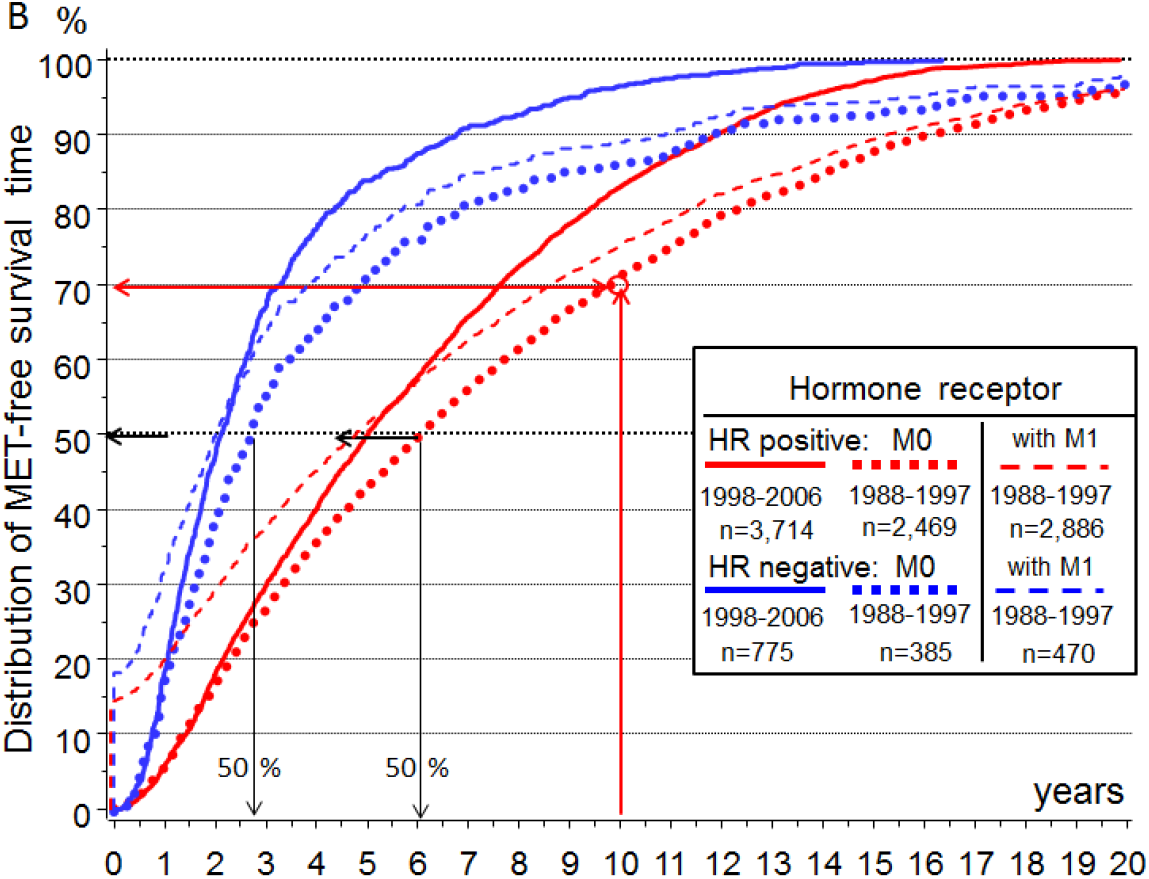
Hormone receptor dependent MET-free survival Distribution functions of the MET-free times for T-N-M0 and HR+ positive (red) and HR- (blue) PTs from the time periods 1988-97 and 1998-2007. Because of the observation period of 20 years the distributions from 1998-2007 reach almost 100% at 20 years. About 30% of the MET appear after 10 years. The dashed lines also show the fraction of T-N-M1 for 1988-97.

**Figure 3C.**
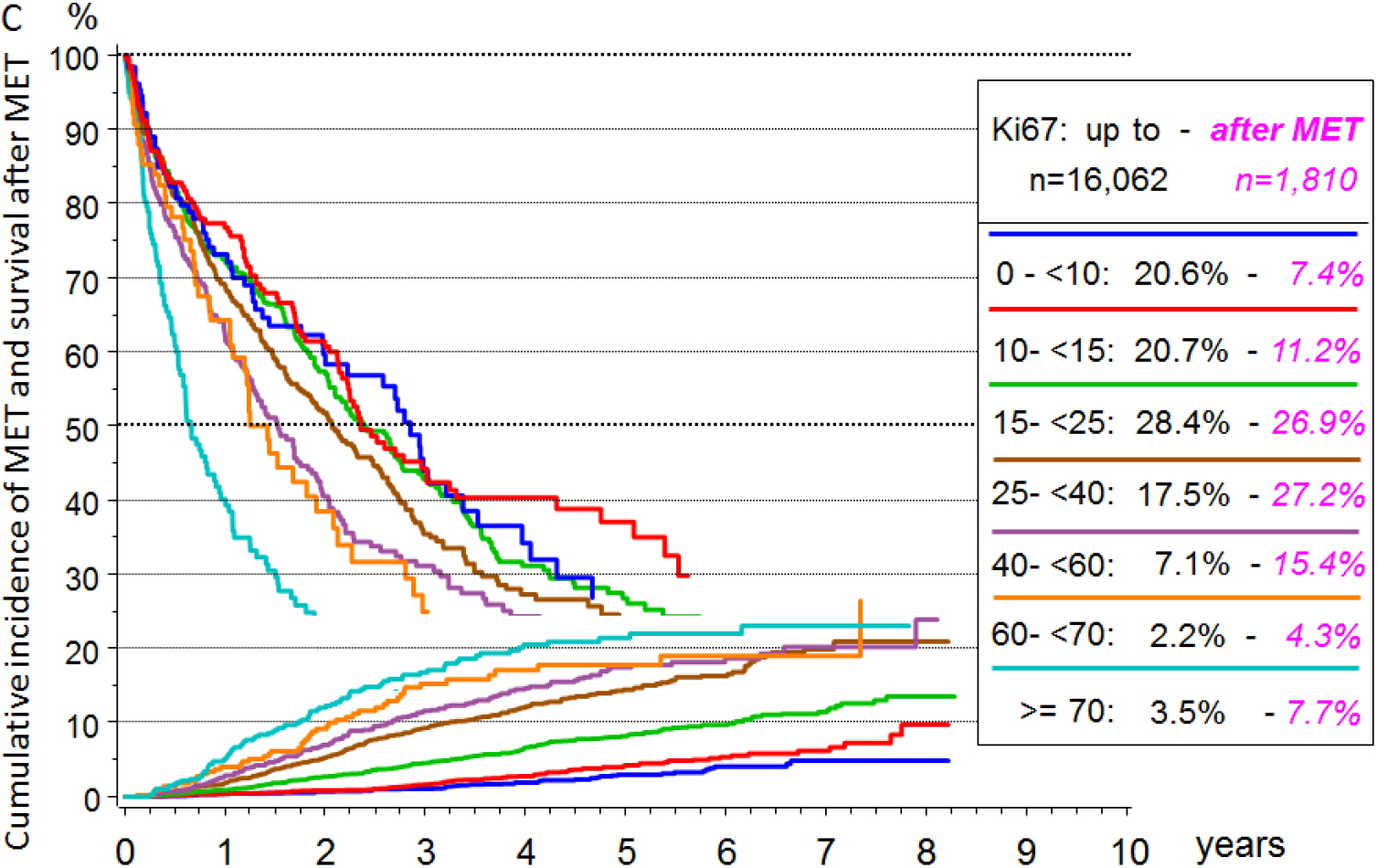
Cumulative incidence of METs and survival after MET depending on KI67. The median survival at T-N-M0 in the event of tumor-related death for the two outer subgroups is approximately 3.1 and 1.5 years. The post MET survival curves are covered at 22%.

**Figure 3D.**
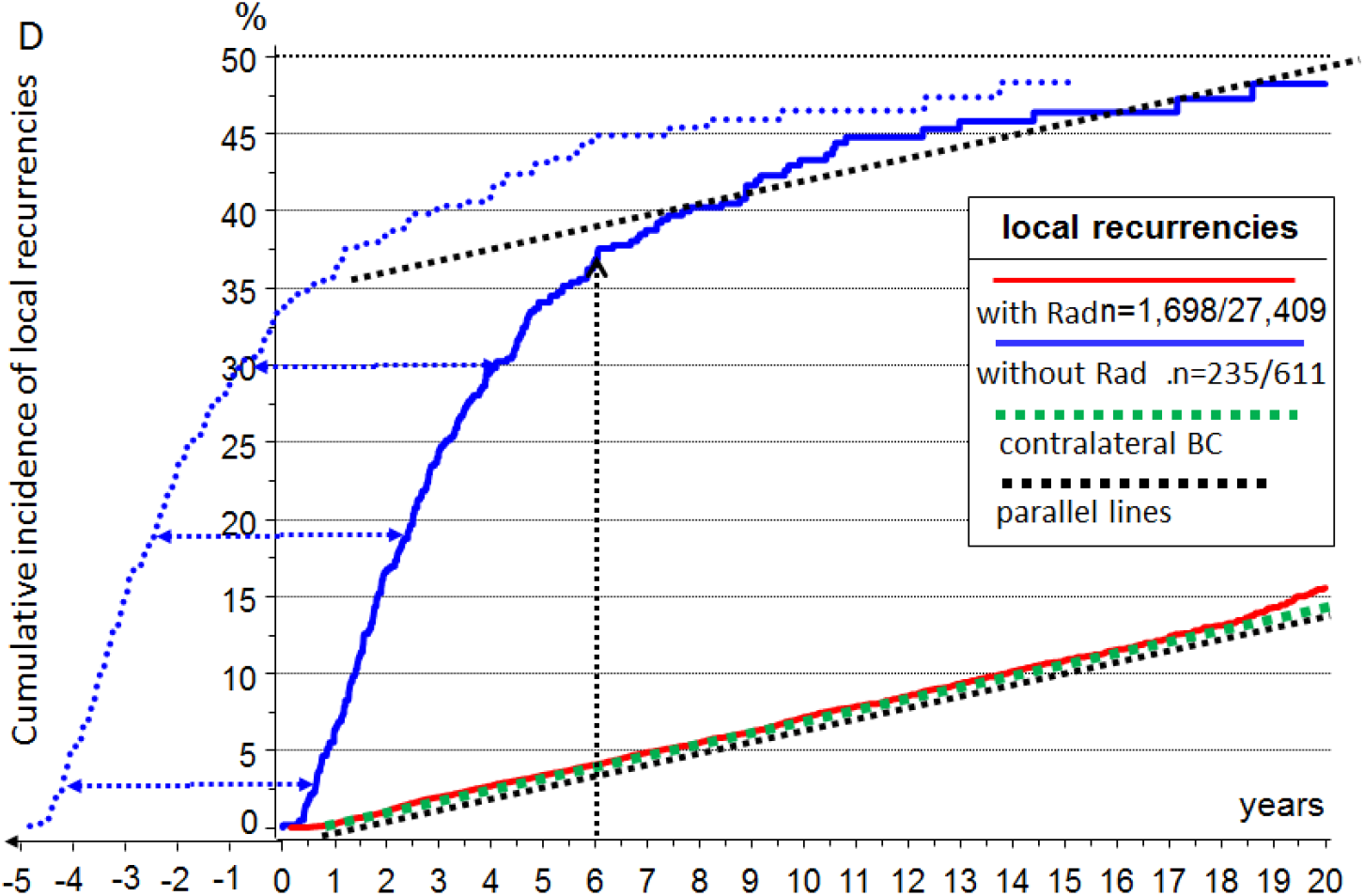
Cumulative incidence of LRs after breast conserving surgery with and without irradiation. Black dotted lines are parallels and show that late LRs increases in both groups comparable to the incidence of contralateral 2ndPT. The dotted blue curve is similar to those for LRs without irradiation and is the fictive initiation of true LR after forwarding the twofold LR-free time of about 26 months.

**Figure 3E.**
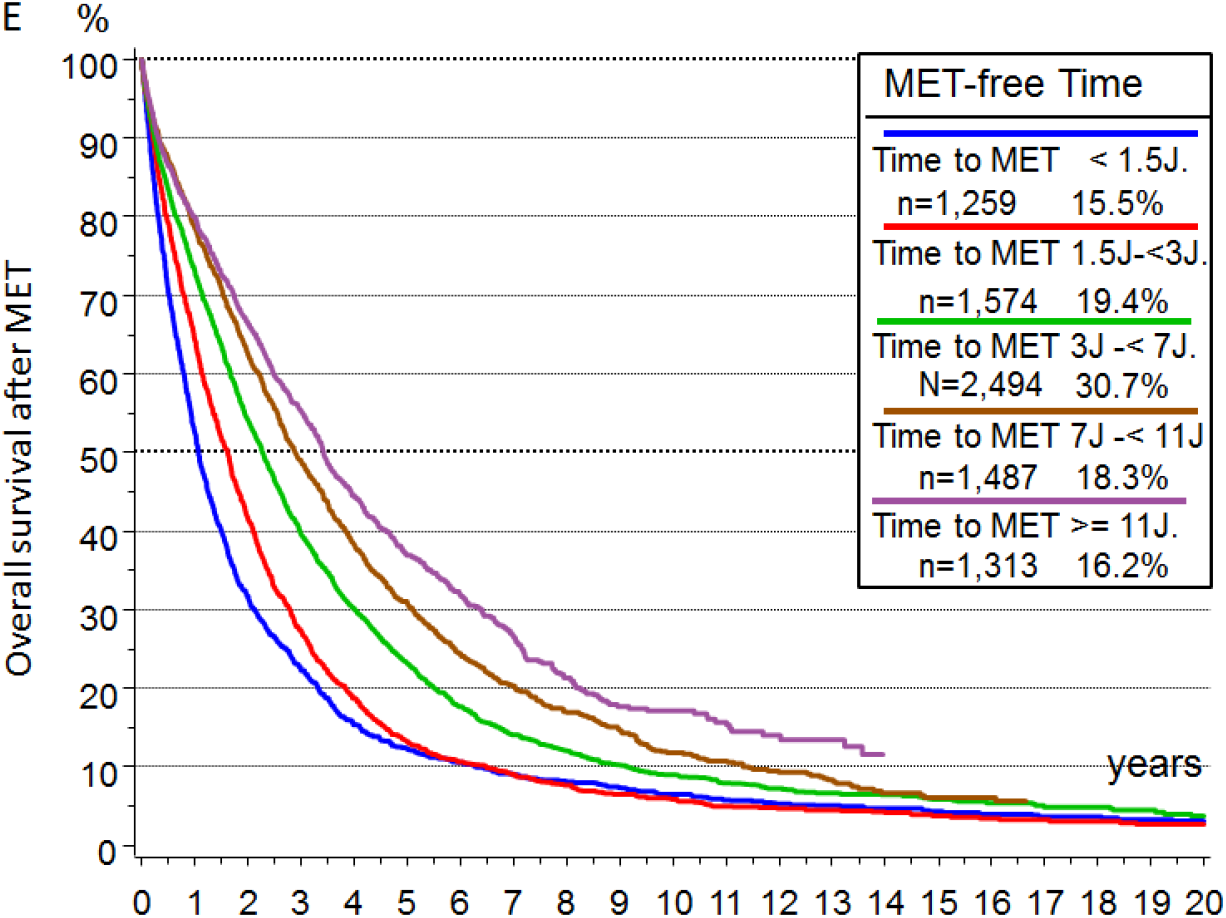
Overall survival from MET of HR+ PTs as a function of MET-free time PTs are diagnosed between 1988-2006 with > 20 years follow-up and n=8,127 METs observed. The differences of PTs of the 1st and 5th Quintile (<1.5, ≥11 ys) are 12.4/36.9% for 5 years survival, 23.9%/45.7% for pT1 and 18.3 / 42.4% for pN0.

**Figure 3F.**
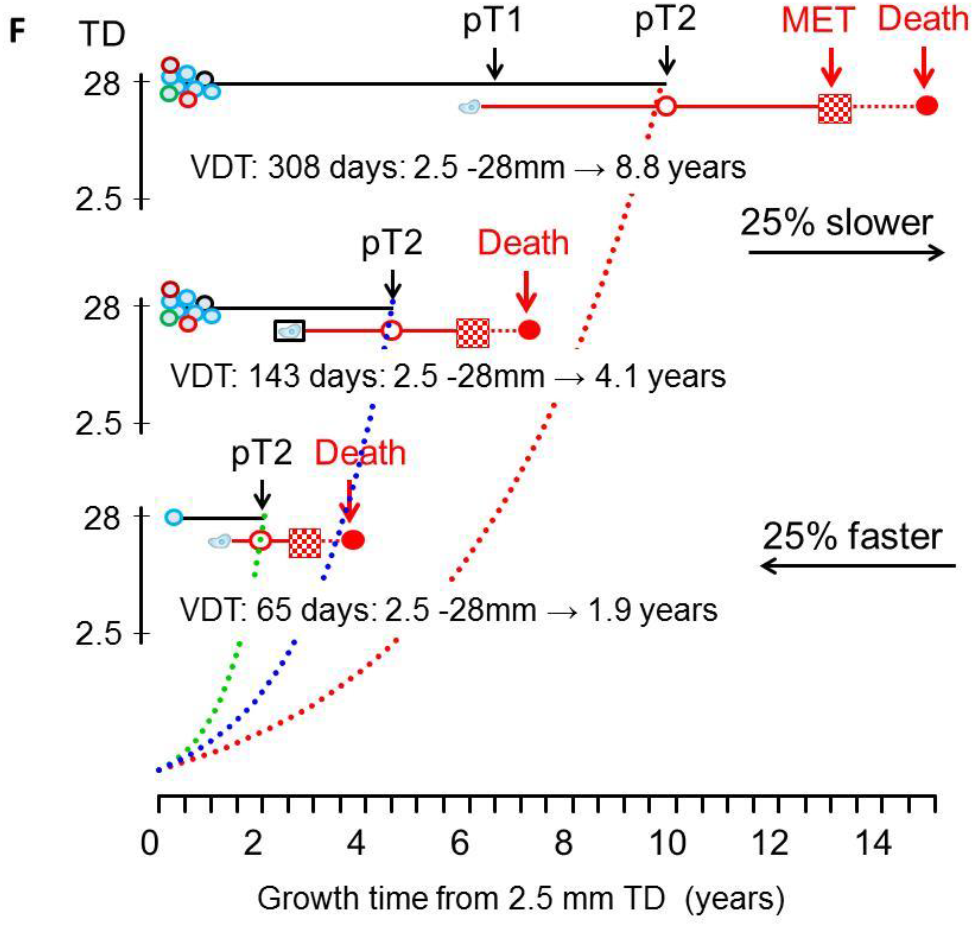
PT/MET growth relation principle Relation of tumor growth of PTs and METs (pT1,pT2), MET-free survival and post MET-survival (red) at different growth rates ^55^

**Figure 3G.**
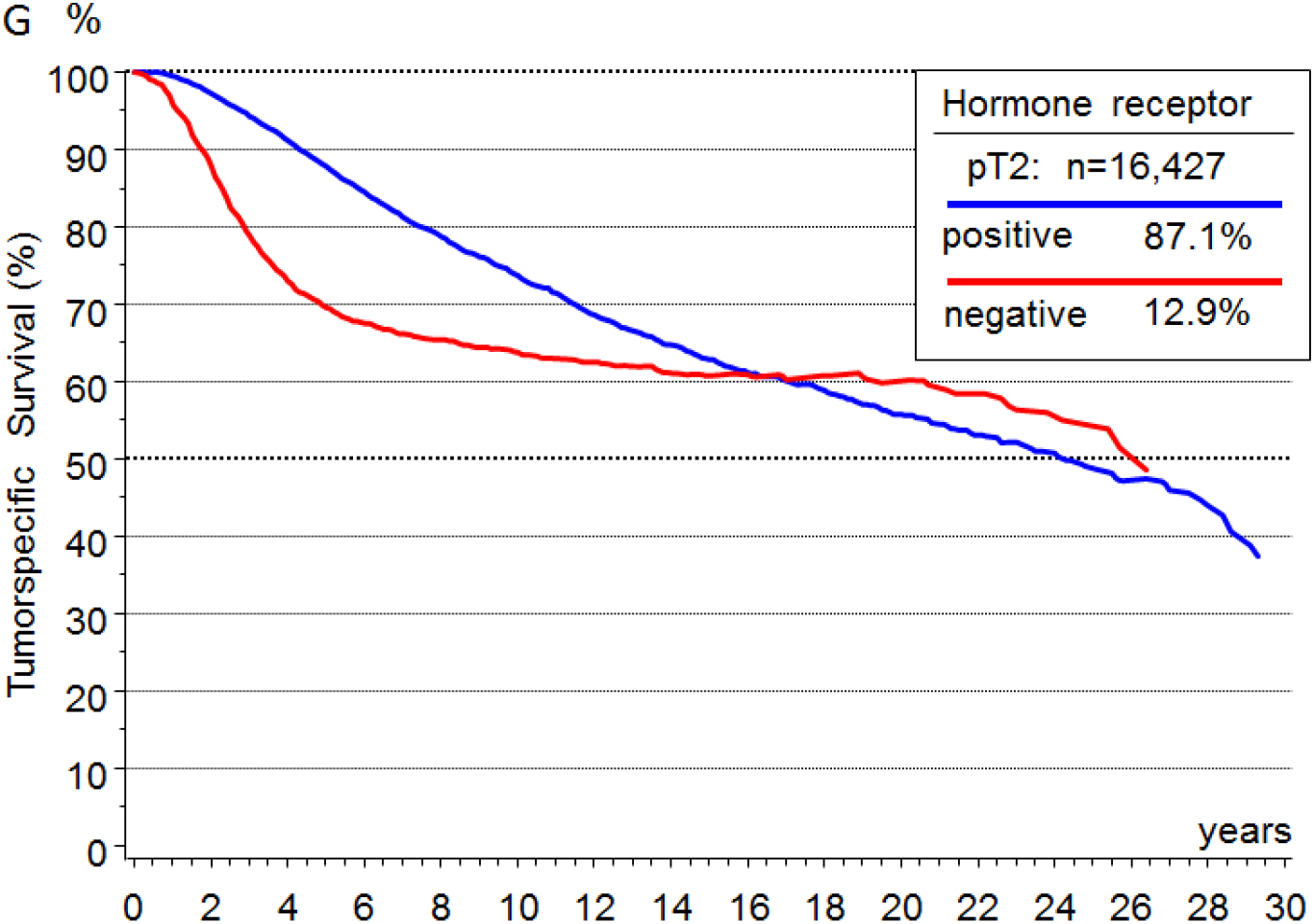
Relative survival of HR+ and HR-PTs.

**Figure 3H.**
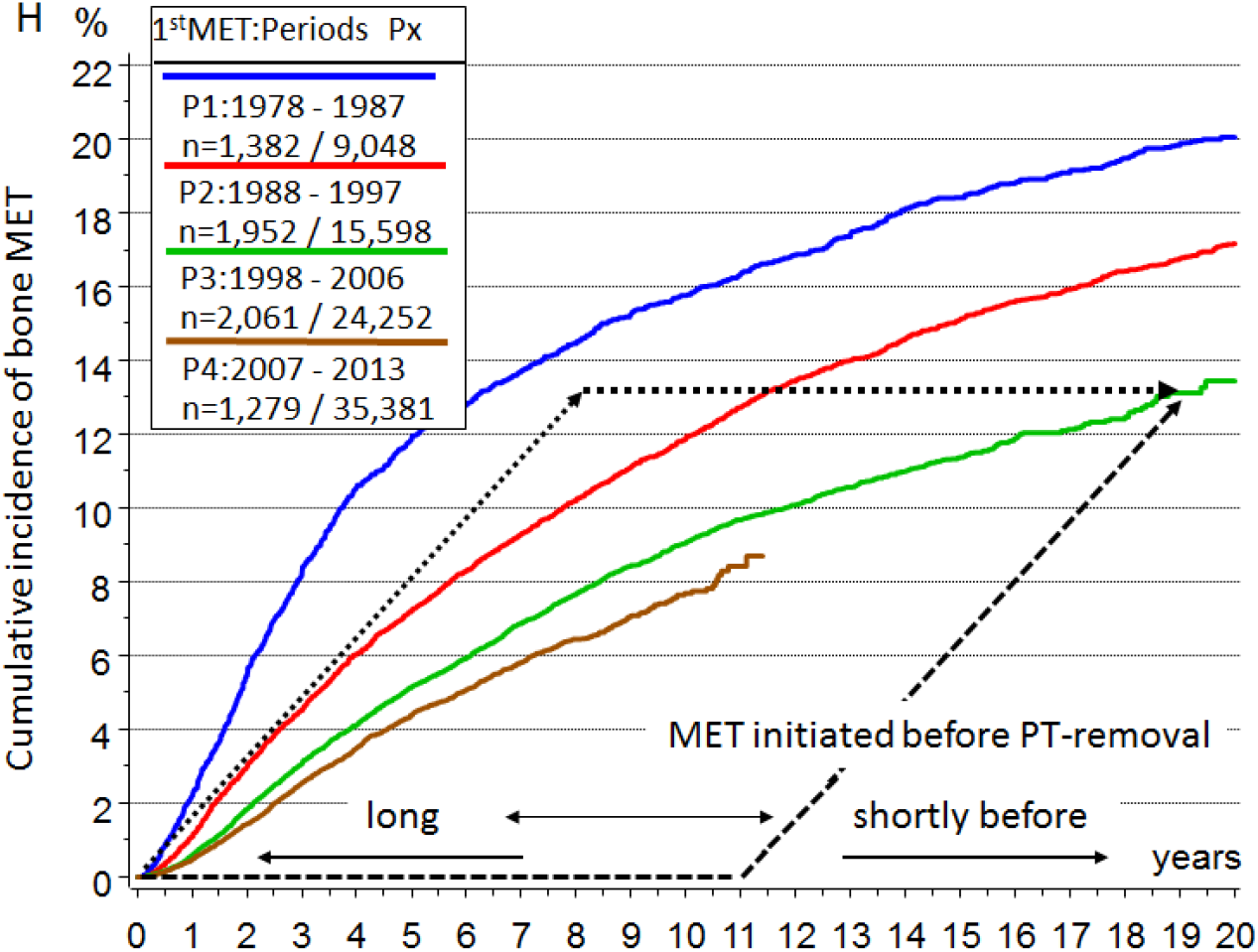
Time trend of bone-MET-free survival. Cumulative incidence of bone METs as a function of 4 time periods since 1978. (Kaplan-Meier method without concurrent risks) The black lines sketch fictitious selective eradication of early (dashed) or late (dottet) initiated METs alternative to green line.

## Notes

### Competing Interest Statement

The authors have declared no competing interest.

### Funding Statement

No external funding was received

### Author Declarations

In the paper, only population-based observational data from a cancer registry for a population of about 4.9 million is used. The registry is regulated by a cancer registry law.

### Summary of Updates

+ The 10 principles and some sections are more precisely formulated due to many suggestions. + the literature was updated, 115 instead of 104

## References

1. Valastyan S, Weinberg RA: Tumor metastasis: molecular insights and evolving paradigms. Cell 147:275–92, 2011

2. Hanahan D, Weinberg RA: Hallmarks of cancer: the next generation. Cell 144:646–74, 2011

3. Yates LR, Knappskog S, Wedge D, et al: Genomic Evolution of Breast Cancer Metastasis and Relapse. Cancer Cell 32:169-184.e7, 2017

4. Lambert AW, Pattabiraman DR, Weinberg RA: Emerging Biological Principles of Metastasis. Cell 168:670–691, 2017

5. Peinado H, Zhang H, Matei IR, et al: Pre-metastatic niches: organ-specific homes for metastases. Nat Rev Cancer 17:302–317, 2017

6. Husemann Y, Geigl JB, Schubert F, et al: Systemic spread is an early step in breast cancer. Cancer Cell 13:58–68, 2008

7. Narod SA, Sopik V: Is invasion a necessary step for metastases in breast cancer? Breast Cancer Res Treat 169:9–23, 2018

8. Butler TP, Gullino PM: Quantitation of cell shedding into efferent blood of mammary adenocarcinoma. Cancer Res 35:512–6, 1975

9. Menyailo ME, Tretyakova MS, Denisov EV: Heterogeneity of Circulating Tumor Cells in Breast Cancer: Identifying Metastatic Seeds. Int J Mol Sci 21, 2020

10. Cresswell GD, Nichol D, Spiteri I, et al: Mapping the breast cancer metastatic cascade onto ctDNA using genetic and epigenetic clonal tracking. Nat Commun 11, 2020

11. Bidard FC, Peeters DJ, Fehm T, et al: Clinical validity of circulating tumour cells in patients with metastatic breast cancer: a pooled analysis of individual patient data. Lancet Oncol 15:406–14, 2014

12. Fisher B, Anderson S, Bryant J, et al: Twenty-year follow-up of a randomized trial comparing total mastectomy, lumpectomy, and lumpectomy plus irradiation for the treatment of invasive breast cancer. N Engl J Med 347:1233–41, 2002

13. Panet-Raymond V, Truong PT, McDonald RE, et al: True recurrence versus new primary: an analysis of ipsilateral breast tumor recurrences after breast-conserving therapy. Int J Radiat Oncol Biol Phys 81:409–17, 2011

14. Whipp E, Beresford M, Sawyer E, et al: True local recurrence rate in the conserved breast after magnetic resonance imaging-targeted radiotherapy. Int J Radiat Oncol Biol Phys 76:984–90, 2010

15. Comen E, Norton L: Self-seeding in cancer. Recent Results Cancer Res 195:13–23, 2012

16. Kim MY, Oskarsson T, Acharyya S, et al: Tumor self-seeding by circulating cancer cells. Cell 139:1315–26, 2009

17. Brierley JD, Gospodarowicz MK, Wittekind C, et al: The TNM Classification of Malignant Tumours 8th ed. Wiley & Sons, 2016

18. Engel J, Weichert W, Jung A, et al: Lymph node infiltration, parallel metastasis and treatment success in breast cancer. Breast 48:1–6, 2019

19. Mamounas EP, Kuehn T, Rutgers EJT, et al: Current approach of the axilla in patients with early-stage breast cancer. Lancet, 2017

20. Munich Cancer Registry: http://www.tumorregister-muenchen.de/en/ (accessed Sept. 20, 2021).

21. Keller L, Pantel K: Unravelling tumour heterogeneity by single-cell profiling of circulating tumour cells. Nat Rev Cancer 19:553–567, 2019

22. Turajlic S, Swanton C: Metastasis as an evolutionary process. Science 352:169–75, 2016

23. Yachida S, Jones S, Bozic I, et al: Distant metastasis occurs late during the genetic evolution of pancreatic cancer. Nature 467:1114–7, 2010

24. Zhang XH, Jin X, Malladi S, et al: Selection of bone metastasis seeds by mesenchymal signals in the primary tumor stroma. Cell 154:1060–73, 2013

25. Bos PD, Zhang XH, Nadal C, et al: Genes that mediate breast cancer metastasis to the brain. Nature 459:1005–9, 2009

26. Weiss L: Comments on hematogenous metastatic patterns in humans as revealed by autopsy. Clin Exp Metastasis 10:191–9, 1992

27. Noone A, Howlader N, Krapcho M, et al: SEER Cancer Statistics Review, 1975-2017 National Cancer Institute. Bethesda, MD,: http://seer.cancer.gov/ x(accessed Sept. 20, 2021).

28. Cuzick J, Sestak I, Cawthorn S, et al: Tamoxifen for prevention of breast cancer: extended long-term follow-up of the IBIS-I breast cancer prevention trial. Lancet Oncol 16:67–75, 2015

29. Gail MH, Brinton LA, Byar DP, et al: Projecting individualized probabilities of developing breast cancer for white females who are being examined annually. J Natl Cancer Inst 81:1879–86, 1989

30. Chaudary MA, Millis RR, Hoskins EO, et al: Bilateral primary breast cancer: a prospective study of disease incidence. Br J Surg 71:711–4, 1984

31. Aguirre-Ghiso JA, Bragado P, Sosa MS: Metastasis Awakening: Targeting dormant cancer. Nat Med 19:276–7, 2013

32. Oskarsson T, Batlle E, Massague J: Metastatic stem cells: sources, niches, and vital pathways. Cell Stem Cell 14:306–21, 2014

33. Meng S, Tripathy D, Frenkel EP, et al: Circulating tumor cells in patients with breast cancer dormancy. Clin Cancer Res 10:8152–62, 2004

34. Bushnell GG, Deshmukh AP, den Hollander P, et al: Breast cancer dormancy: need for clinically relevant models to address current gaps in knowledge. NPJ Breast Cancer 7:66, 2021

35. Werner S, Heidrich I, Pantel K: Clinical management and biology of tumor dormancy in breast cancer. Semin Cancer Biol, 2021

36. Janni W, Vogl FD, Wiedswang G, et al: Persistence of disseminated tumor cells in the bone marrow of breast cancer patients predicts increased risk for relapse--a European pooled analysis. Clin Cancer Res 17:2967–76, 2011

37. Early Breast Cancer Trialists’ Collaborative Group: Systemic treatment of early breast cancer by hormonal, cytotoxic, or immune therapy. 133 randomised trials involving 31,000 recurrences and 24,000 deaths among 75,000 women. Early Breast Cancer Trialists’ Collaborative Group. Lancet 339:71–85, 1992

38. Hunter KW, Amin R, Deasy S, et al: Genetic insights into the morass of metastatic heterogeneity. Nat Rev Cancer 18:211–223, 2018

39. Early Breast Cancer Trialists’ Collaborative Group: Effects of radiotherapy and of differences in the extent of surgery for early breast cancer on local recurrence and 15-year survival: an overview of the randomised trials. Lancet 366:2087–106, 2005

40. Early Breast Cancer Trialists’ Collaborative Group: Effect of radiotherapy after breast-conserving surgery on 10-year recurrence and 15-year breast cancer death: meta-analysis of individual patient data for 10 801 women in 17 randomised trials. Lancet 378:1707–16, 2011

41. Ullah I, Karthik GM, Alkodsi A, et al: Evolutionary history of metastatic breast cancer reveals minimal seeding from axillary lymph nodes. J Clin Invest 128:1355–1370, 2018

42. Cady B: Lymph node metastases. Indicators, but not governors of survival. Arch Surg 119:1067–72, 1984

43. Engel J, Emeny RT, Holzel D: Positive lymph nodes do not metastasize. Cancer Metastasis Rev 31:235–246, 2012

44. Giuliano AE, Ballman KV, McCall L, et al: Effect of Axillary Dissection vs No Axillary Dissection on 10-Year Overall Survival Among Women With Invasive Breast Cancer and Sentinel Node Metastasis: The ACOSOG Z0011 (Alliance) Randomized Clinical Trial. Jama 318:918–926, 2017

45. Engel J, Lebeau A, Sauer H, et al: Are we wasting our time with the sentinel technique? Fifteen reasons to stop axilla dissection. Breast 15:452–5, 2006

46. Halsted W: The results of operations for the cure of the cancer of the breast performed at the Johns Hopkins Hospital from June 1889 to January 1894. Arch Surgery 20:497, 1894

47. Crile G: Excision of cancer of the head and neck. JAMA 47:1780–1786, 1906

48. Pereira ER, Kedrin D, Seano G, et al: Lymph node metastases can invade local blood vessels, exit the node, and colonize distant organs in mice. Science 359:1403–1407, 2018

49. Cristofanilli M, Budd GT, Ellis MJ, et al: Circulating tumor cells, disease progression, and survival in metastatic breast cancer. N Engl J Med 351:781–91, 2004

50. Mamounas EP, Mitchell MP, Woodward WA: Molecular Predictive and Prognostic Markers in Locoregional Management. J Clin Oncol 38:2310–2320, 2020

51. Gundem G, Van Loo P, Kremeyer B, et al: The evolutionary history of lethal metastatic prostate cancer. Nature 520:353–357, 2015

52. Ramaswamy S, Ross KN, Lander ES, et al: A molecular signature of metastasis in primary solid tumors. Nat Genet 33:49–54, 2003

53. Hölzel D, Eckel R, Emeny R, et al: Distant metastases do not metastasize. Cancer Metastasis Rev 29:737–750, 2010

54. Spratt JA, von Fournier D, Spratt JS, et al: Decelerating growth and human breast cancer. Cancer 71:2013–9, 1993

55. Weedon-Fekjaer H, Lindqvist BH, Vatten LJ, et al: Breast cancer tumor growth estimated through mammography screening data. Breast Cancer Res 10:R41, 2008

56. Houssami N, Hunter K: The epidemiology, radiology and biological characteristics of interval breast cancers in population mammography screening. NPJ Breast Cancer 3:12, 2017

57. Zhang S, Ding Y, Zhou Q, et al: Correlation Factors Analysis of Breast Cancer Tumor Volume Doubling Time Measured by 3D-Ultrasound. Med Sci Monit 23:3147–3153, 2017

58. Cuzick J: Preventive therapy for cancer. Lancet Oncol 18:e472–e482, 2017

59. Chlebowski RT, Kuller LH, Prentice RL, et al: Breast cancer after use of estrogen plus progestin in postmenopausal women. N Engl J Med 360:573–87, 2009

60. Chlebowski RT, Hendrix SL, Langer RD, et al: Influence of estrogen plus progestin on breast cancer and mammography in healthy postmenopausal women: the Women’s Health Initiative Randomized Trial. JAMA 289:3243–53, 2003

61. Engel J, Schubert-Fritschle G, Hölzel D: Hormone replacement therapy and elevated breast cancer risk: An artifact of growth acceleration? MEDRXIV/2020/050708, 2020

62. Bertucci F, Ng CKY, Patsouris A, et al: Genomic characterization of metastatic breast cancers. Nature 569:560–564, 2019

63. Pan H, Gray R, Braybrooke J, et al: 20-Year Risks of Breast-Cancer Recurrence after Stopping Endocrine Therapy at 5 Years. N Engl J Med 377:1836–1846, 2017

64. Hölzel D, Eckel R, Bauerfeind I, et al: Improved systemic treatment for early breast cancer improves cure rates, modifies metastatic pattern and shortens post-metastatic survival: 35-year results from the Munich Cancer Registry. J Cancer Res Clin Oncol 143:1701–12, 2017

65. McGrath S, Antonucci J, Goldstein N, et al: Long-term patterns of in-breast failure in patients with early stage breast cancer treated with breast-conserving therapy: a molecular based clonality evaluation. Am J Clin Oncol 33:17–22, 2010

66. Smith TE, Lee D, Turner BC, et al: True recurrence vs. new primary ipsilateral breast tumor relapse: an analysis of clinical and pathologic differences and their implications in natural history, prognoses, and therapeutic management. Int J Radiat Oncol Biol Phys 48:1281–9, 2000

67. Colleoni M, Rotmensz N, Peruzzotti G, et al: Size of breast cancer metastases in axillary lymph nodes: clinical relevance of minimal lymph node involvement. J Clin Oncol 23:1379–89, 2005

68. de Boer M, van Deurzen CH, van Dijck JA, et al: Micrometastases or isolated tumor cells and the outcome of breast cancer. N Engl J Med 361:653–63, 2009

69. Pedersen L, Gunnarsdottir KA, Rasmussen BB, et al: The prognostic influence of multifocality in breast cancer patients. Breast 13:188–93, 2004

70. O’Daly BJ, Sweeney KJ, Ridgway PF, et al: The accuracy of combined versus largest diameter in staging multifocal breast cancer. J Am Coll Surg 204:282–5, 2007

71. Hölzel D, Eckel R, Bauerfeind I, et al: Survival of de novo stage IV breast cancer patients over three decades. J Cancer Res Clin Oncol 143:509–19, 2017

72. Colleoni M, Sun Z, Price KN, et al: Annual Hazard Rates of Recurrence for Breast Cancer During 24 Years of Follow-Up: Results From the International Breast Cancer Study Group Trials I to V. J Clin Oncol 34:927–35, 2016

73. Cortazar P, Zhang L, Untch M, et al: Pathological complete response and long-term clinical benefit in breast cancer: the CTNeoBC pooled analysis. Lancet 384:164–72, 2014

74. Robidoux A, Tang G, Rastogi P, et al: Lapatinib as a component of neoadjuvant therapy for HER2-positive operable breast cancer (NSABP protocol B-41): an open-label, randomised phase 3 trial. Lancet Oncol 14:1183–92, 2013

75. von Minckwitz G, Untch M, Blohmer JU, et al: Definition and impact of pathologic complete response on prognosis after neoadjuvant chemotherapy in various intrinsic breast cancer subtypes. J Clin Oncol 30:1796–804, 2012

76. Fontein DB, Charehbili A, Nortier JW, et al: Efficacy of six month neoadjuvant endocrine therapy in postmenopausal, hormone receptor-positive breast cancer patients--a phase II trial. Eur J Cancer 50:2190–200, 2014

77. Fentiman IS, Christiaens MR, Paridaens R, et al: Treatment of operable breast cancer in the elderly: a randomised clinical trial EORTC 10851 comparing tamoxifen alone with modified radical mastectomy. Eur J Cancer 39:309–16, 2003

78. Spring LM, Gupta A, Reynolds KL, et al: Neoadjuvant Endocrine Therapy for Estrogen Receptor-Positive Breast Cancer: A Systematic Review and Meta-analysis. JAMA Oncol 2:1477–1486, 2016

79. Ellis MJ, Suman VJ, Hoog J, et al: Ki67 Proliferation Index as a Tool for Chemotherapy Decisions During and After Neoadjuvant Aromatase Inhibitor Treatment of Breast Cancer: Results From the American College of Surgeons Oncology Group Z1031 Trial (Alliance). J Clin Oncol 35:1061–1069, 2017

80. Fisher B, Costantino JP, Wickerham DL, et al: Tamoxifen for the prevention of breast cancer: current status of the National Surgical Adjuvant Breast and Bowel Project P-1 study. J Natl Cancer Inst 97:1652–62, 2005

81. Cuzick J, Sestak I, Forbes JF, et al: Use of anastrozole for breast cancer prevention (IBIS-II): long-term results of a randomised controlled trial. Lancet 395:117–22, 2020

82. Early Breast Cancer Trialists’ Collaborative Group: Tamoxifen for early breast cancer: an overview of the randomised trials. Early Breast Cancer Trialists’ Collaborative Group. Lancet 351:1451–67, 1998

83. Early Breast Cancer Trialists’ Collaborative Group: Relevance of breast cancer hormone receptors and other factors to the efficacy of adjuvant tamoxifen: patient-level meta-analysis of randomised trials. Lancet 378:771–84, 2011

84. Early Breast Cancer Trialists’ Collaborative Group: Comparisons between different polychemotherapy regimens for early breast cancer: meta-analyses of long-term outcome among 100,000 women in 123 randomised trials. Lancet 379:432–44, 2012

85. Cameron D, Piccart-Gebhart MJ, Gelber RD, et al: 11 years’ follow-up of trastuzumab after adjuvant chemotherapy in HER2-positive early breast cancer: final analysis of the HERceptin Adjuvant (HERA) trial. Lancet 389:1195–1205, 2017

86. Welch HG, Prorok PC, O’Malley AJ, et al: Breast-Cancer Tumor Size, Overdiagnosis, and Mammography Screening Effectiveness. N Engl J Med 375:1438–1447, 2016

87. Jurrius P, Green T, Garmo H, et al: Invasive breast cancer over four decades reveals persisting poor metastatic outcomes in treatment resistant subgroup - the “ATRESS” phenomenon. Breast 50:39–48, 2020

88. Hanna TP, King WD, Thibodeau S, et al: Mortality due to cancer treatment delay: systematic review and meta-analysis. Bmj 371:m4087, 2020

89. Delozier T, Switsers O, Genot JY, et al: Delayed adjuvant tamoxifen: ten-year results of a collaborative randomized controlled trial in early breast cancer (TAM-02 trial). Ann Oncol 11:515–9, 2000

90. Veronesi A, Miolo G, Magri MD, et al: Late tamoxifen in patients previously operated for breast cancer without postoperative tamoxifen: 5-year results of a single institution randomised study. BMC Cancer 10:205, 2010

91. Kim H, Kim CY, Park KH, et al: Clonality analysis of multifocal ipsilateral breast carcinomas using X-chromosome inactivation patterns. Hum Pathol 78:106–114, 2018

92. Mamtani A, Patil S, Stempel M, et al: Axillary Micrometastases and Isolated Tumor Cells Are Not an Indication for Post-Mastectomy Radiotherapy in Stage 1 and 2 Breast Cancer. Ann Surg Oncol 24:2182–8, 2017

93. Razavi P, Chang MT, Xu G, et al: The Genomic Landscape of Endocrine-Resistant Advanced Breast Cancers. Cancer Cell 34:427-438.e6, 2018

94. Goldhirsch A, Gelber RD, Piccart-Gebhart MJ, et al: 2 years versus 1 year of adjuvant trastuzumab for HER2-positive breast cancer (HERA): an open-label, randomised controlled trial. Lancet 382:1021–8, 2013

95. Peto R: Five years of tamoxifen--or more? J Natl Cancer Inst 88:1791–3, 1996

96. Smith IE, Yeo B, Schiavon G: The optimal duration and selection of adjuvant endocrine therapy for breast cancer: how long is enough? Am Soc Clin Oncol Educ Book:e16–24, 2014

97. Pivot X, Romieu G, Debled M, et al: 6 months versus 12 months of adjuvant trastuzumab for patients with HER2-positive early breast cancer (PHARE): a randomised phase 3 trial. Lancet Oncol 14:741–8, 2013

98. Petrelli F, Rulli E, Labianca R, et al: Overall survival with 3 or 6 months of adjuvant chemotherapy in Italian TOSCA Phase 3 Randomized Trial. Ann Oncol, 2020

99. Gnant M, Sestak I, Filipits M, et al: Identifying clinically relevant prognostic subgroups of postmenopausal women with node-positive hormone receptor-positive early-stage breast cancer treated with endocrine therapy: a combined analysis of ABCSG-8 and ATAC using the PAM50 risk of recurrence score and intrinsic subtype. Ann Oncol 26:1685–91, 2015

100. van ‘t Veer LJ, Yau C, Yu NY, et al: Tamoxifen therapy benefit for patients with 70-gene signature high and low risk. Breast Cancer Res Treat 166:593–601, 2017

101. Zhang L, Hsieh MC, Petkov V, et al: Trend and survival benefit of Oncotype DX use among female hormone receptor-positive breast cancer patients in 17 SEER registries, 2004-2015. Breast Cancer Res Treat 180:491–501, 2020

102. Anandan A, Sharifi M, O’Regan R: Molecular Assays to Determine Optimal Duration of Adjuvant Endocrine Therapy in Breast Cancer. Curr Treat Options Oncol 21:84, 2020

103. Davies C, Pan H, Godwin J, et al: Long-term effects of continuing adjuvant tamoxifen to 10 years versus stopping at 5 years after diagnosis of oestrogen receptor-positive breast cancer: ATLAS, a randomised trial. Lancet 381:805–16, 2013

104. Curigliano G, Burstein HJ E PW, et al: De-escalating and escalating treatments for early-stage breast cancer: the St. Gallen International Expert Consensus Conference on the Primary Therapy of Early Breast Cancer 2017. Ann Oncol 28:1700–1712, 2017

105. Burstein HJ, Lacchetti C, Anderson H, et al: Adjuvant Endocrine Therapy for Women With Hormone Receptor-Positive Breast Cancer: ASCO Clinical Practice Guideline Focused Update. J Clin Oncol 37:423–438, 2019

106. Engel J, Schubert-Fritschle G, Emeny R, et al: Breast cancer: Are long-term and intermittent endocrine therapies equally effective? J Cancer Res Clin Oncol, 2020

107. Minchinton AI, Tannock IF: Drug penetration in solid tumours. Nat Rev Cancer 6:583–92, 2006

108. Conte P, Frassoldati A, Bisagni G, et al: 9 weeks vs 1 year adjuvant trastuzumab in combination with chemotherapy: final results of the phase III randomized Short-HER study. Ann Oncol, 2018

109. Holohan C, Van Schaeybroeck S, Longley DB, et al: Cancer drug resistance: an evolving paradigm. Nat Rev Cancer 13:714–26, 2013

110. Jeselsohn R, Buchwalter G, De Angelis C, et al: ESR1 mutations-a mechanism for acquired endocrine resistance in breast cancer. Nat Rev Clin Oncol 12:573–83, 2015

111. Musgrove EA, Sutherland RL: Biological determinants of endocrine resistance in breast cancer. Nat Rev Cancer 9:631–43, 2009

112. Vivot A, Jacot J, Zeitoun JD, et al: Clinical benefit, price and approval characteristics of FDA-approved new drugs for treating advanced solid cancer, 2000-2015. Ann Oncol 28:1111–1116, 2017

113. Altrock PM, Liu LL, Michor F: The mathematics of cancer: integrating quantitative models. Nat Rev Cancer 15:730–45, 2015

114. Hershman DL, Wright JD: Comparative effectiveness research in oncology methodology: observational data. J Clin Oncol 30:4215–22, 2012

115. Concato J, Lawler EV, Lew RA, et al: Observational methods in comparative effectiveness research. Am J Med 123:e16–23, 2010

